# The Impact of Opioid, Opioid Agonist Therapy, and Cannabis Exposure on Fetal Growth: A Population-Based Cohort Study

**DOI:** 10.64898/2026.09.10.26362743

**Authors:** Jessica Pudwell, Kira King, Wenbin Li, Maria Velez, Shannon Bainbridge, Laura Gaudet

## Abstract

**Objectives:** The primary objective is to determine the extent to which fetal growth profiles in opioid and opioid agonist (OAT) exposed pregnancies are influenced by cannabis exposure. Secondary objectives are to examine differences in fetal and neonatal morbidity and mortality.

**Design:** Population-based cohort study.

**Setting:** Ontario, Canada, in a public healthcare system.

**Participants:** All live/stillborn births between April 1^st^, 2013, and March 31^st^, 2021.

**Exposures:** Substance exposure in pregnancy, including opioids, opioid agonist therapy, cannabis, and nicotine.

**Main Outcome Measures:** Primary outcome measures included incidence of small for gestational age (<3^rd^ and <10^th^ percentile for sex) and intrauterine growth restriction. Secondary outcome measures included incidence of stillbirth, severe neonatal morbidity (SNM), neonatal mortality, and neonatal abstinence syndrome (NAS).

**Results:** 959,731 births are included with exposures classified as 864,508 (88.0%) no substance, 73,815 (7.7%) nicotine, 23,003 (2.4%) cannabis, 7,694 (0.8%) opioid, and 5,353 (0.6%) OAT. Cannabis co-exposure was reported in 1 out of every 6 opioid and/or OAT exposed births. The observed proportions of SGA and IUGR were approximately doubled across substance-exposed groups compared with the no substance exposure group. In adjusted analyses, cannabis exposure alone was associated with an 84% increased risk of IUGR, compared with 14% for opioid exposure alone and 60% for OAT exposure alone. SNM was observed in 7.2% of no substance exposure neonates, compared to 13.9% and 14.2% of opioid and OAT exposed neonates, respectively. NAS was diagnosed in 70.6% of all OAT exposed neonates, compared to 34.3% of all opioid exposed neonates. Risks for all outcomes across all cannabis co-exposure groups were significantly elevated relative to the no substance exposure group, with estimates ranging from 43% to 107% increased risk.

**Conclusions:** Prenatal co-exposure to opioid and/or OAT along with cannabis appears to place infants at the highest risk. Isolated cannabis use has similar, if not more significant, impacts on fetal growth as opioid and/or OAT use. This information may serve as an important starting point in the design of effective harm reduction strategies in this patient population.

**Tweetable abstract:** Prenatal co-exposure to cannabis and opioid and/or opioid agonist therapy (OAT) use places infants at increased risk for impaired fetal growth and severe neonatal morbidity. Isolated cannabis use in pregnancy has similar, if not greater, impacts on fetal growth than opioid and/or OAT use.

**Summary Boxes:** *Section 1: What is already known on this topic:* Across North America, rates of opioid exposure during pregnancy range from 1.1-42%. Multi-substance use is common, with concurrent cannabis exposure estimated at 36-75% among opioid users. Opioid use in pregnancy is associated with altered fetal growth, including low birthweight and small for gestational age (SGA) infants. Cannabis use in pregnancy is likewise associated with compromised fetal and neonatal growth, along with neurocognitive developmental deficits in offspring. Despite these known risks in isolation, little is known about the impact of co-use, which is concerning given the high rates of concurrent substance exposure.

*Section 2: What this study adds:* This study demonstrates that babies exposed to cannabis, opioids and/or OAT in pregnancy are more likely to experience complications, including failure to reach ideal growth potential, stillbirth, difficulties adapting to life after birth, readmission to hospital within 42 days of delivery, and neonatal death. Prenatal cannabis co-exposure with opioid and/or OAT places infants at the highest risk. Isolated cannabis use has similar, if not greater, impacts on fetal growth than isolated opioid or OAT use, significantly increasing the risks of impaired fetal growth (including fetal growth restriction (FGR) and SGA) and severe neonatal morbidity (SNM).

## Introduction

Opioid use during pregnancy remains a major public health concern within the broader opioid epidemic (1–5). Across North America, prevalence estimates vary considerably, ranging from 1.1 to 42% (6–10), reflecting differences in population characteristics, surveillance methods, and definitions of exposures (11, 12). Multi-substance use is common in this population; among pregnant individuals using opioids, 36-75% also report cannabis use (13–16). Given these high rates of co-exposure, understanding the combined effects of these substances, rather than considering each in isolation, is increasingly important.

Opioid use in pregnancy is associated with neonatal abstinence syndrome (NAS), a withdrawal syndrome in newborns whose incidence has risen sharply in incidence over recent decades (17, 18). Beyond withdrawal, opioid exposure is associated with altered fetal growth profiles, including increased rates of low birthweight and small for gestational age (SGA) infants (8, 9). These fetal growth impairments carry important implications for perinatal and long-term health, including increased risks of preterm birth, stillbirth, and adverse developmental and cardiometabolic outcomes later in life (19–24). Opioid agonist therapy (OAT), most commonly with long-acting opioid agonists such as methadone or buprenorphine, is the recommended first-line treatment for opioid use disorder in pregnancy and is central to clinical harm-reduction strategies. OAT stabilizes maternal opioid levels, prevents withdrawal, reduces cravings and illicit opioid use, and is associated with improved maternal and fetal outcomes compared with ongoing untreated opioid use or withdrawal (25–27). However, OAT-exposed infants remain at increased risk of adverse outcomes, including preterm birth, impaired fetal growth, and neonatal abstinence syndrome, compared with unexposed populations (28). Outcomes may also differ by OAT regimen, with growing evidence suggesting lower risks of preterm birth, low birthweight, SGA, and neonatal withdrawal among buprenorphine-exposed infants compared with those exposed to methadone, highlighting the need for more detailed evaluation (27, 29, 30).

Cannabis exposure in pregnancy has also been linked to impaired fetal growth, independent of opioid use, with dose- and timing-dependent reductions in birthweight, including a reported 244g decrease with continuous use and a 156g decrease when cannabis use ceases in the first half of pregnancy (31). Prenatal cannabis exposure has also been associated with impaired fetal and neonatal growth, as well as alterations in offspring neurodevelopment, particularly in attention, behavioural regulation, and executive functioning (32–34). Despite these established risks associated with opioid, OAT, and cannabis exposures individually, little is known about the impact of co-use, despite its high prevalence. Whether cannabis modifies or augments opioid- or OAT-related risks to fetal growth and neonatal health remains poorly understood. Clarifying whether cannabis co-exposure adds to the risks associated with opioid or OAT use may help inform more targeted counselling and harm-reduction strategies during pregnancy. To address this gap, the primary objective of this study was therefore to determine the extent to which cannabis co-exposure influences fetal growth in opioid and OAT-exposed pregnancies, with secondary objectives examining fetal and neonatal morbidity and mortality.

## Methods

### Study Design and Population

This population-based pregnancy cohort study included pregnancies resulting in a live birth or stillbirth in an Ontario (Canada) hospital between April 1^st^, 2013, and March 31^st^, 2021. Eligible pregnancies met all of the following criteria: maternal age 16-50 years at delivery; ≥ 2 years of Ontario Health Insurance Plan (OHIP) eligibility prior to the estimated date of conception; and gestational age ≥20 weeks at delivery. Ethical approval was obtained from the Queen’s University Health Sciences and Affiliated Teaching Hospitals Research Ethics Board (HSREB File #6040679).

### Data Sources

*ICES data environment* – Analyses were conducted at ICES (www.ices.on.ca), an independent research institute authorized under Ontario’s health information privacy legislation to collect and analyze health care and demographic data without individual consent for health system evaluation.

*Primary perinatal dataset (BORN Ontario)* – Exposure information and pregnancy outcomes were obtained from the Better Outcomes Registry & Network (BORN) Ontario (www.bornontario.ca), which captures > 99% of hospital births in the province and has been validated for completeness and accuracy (35).

*Linked administrative databases* - Maternal demographics, health conditions, and related pregnancy-related data were obtained through linkages to multiple administrative datasets housed at ICES, including the Canadian Institute for Health Information Discharge Abstract Database (CIHI DAD), National Ambulatory Care Reporting System (NACRS), OHIP Claims Database, the Ontario Mental Health Reporting System (OMHRS), the Registered Persons Database (RPDB), the Postal Code Conversion File (PCCF), Linked Delivering Mothers and Newborns (MOMBABY), Ontario Hypertension Dataset (HYPER), and Ontario Diabetes Dataset (ODD). The specific contributions of each dataset and variable definitions are detailed in **Appendix 1** and **Appendix 2**.

### Exposure Classification

Substance exposures in pregnancy were based on maternal self-report and provider documentation in the BORN database. Exposures were captured at any point in pregnancy as recorded in BORN. Participants were classified according to reported use of: Opioids (prescribed or illicit); OAT (e.g., methadone, buprenorphine/Subutex); cannabis; nicotine; and other substances (e.g., cocaine, amphetamines, inhalants, hallucinogens). Combinations of opioid, OAT, and cannabis exposure were used to generate mutually exclusive exposure groups; participants in these groups could also have nicotine exposure. Nicotine and other substance exposures were accounted for separately in adjusted analyses. In BORN, some exposure fields include “unknown” options. As detailed in **Appendix 2**, unknown values were recoded according to prespecified rules (generally grouped with “no” exposure unless stated otherwise). Nicotine and “other substances” exposures were not primary exposures of interest but were accounted for in adjusted analyses. Participants with no reported use of any of these substances formed the reference group.

### Outcomes

Primary outcomes were SGA, defined as birthweight below the 3rd or 10th percentile for gestational age and sex using Canadian reference standards (36), and intrauterine growth restriction (IUGR), based on clinician-documented diagnosis in BORN.

Secondary outcomes include stillbirth, severe neonatal morbidity (SNM), neonatal mortality, and neonatal abstinence syndrome (NAS). Stillbirth, neonatal mortality, and NAS were identified using BORN, MOMBABY, and/or RPDB linkages according to standard definitions. SNM was assessed using a validated composite of major neonatal diagnoses and interventions within 28 days of life (37). The full list of ICD-10-CA/CCI codes used to construct the composite is contained in **Appendix 2**. For multiple gestations, outcomes were modelled per pregnancy; if any infant met the definition, the pregnancy was coded as positive.

### Covariates

Covariates were selected a priori based on their established associations with fetal growth and neonatal outcomes. Detailed coding definitions and data sources for all covariates are provided in **Appendix 2**.

Demographic characteristics included maternal age at delivery, parity, neighborhood income quintile as a measure of socioeconomic status, and rural versus urban residence based on the Rurality Index of Ontario. Pre-pregnancy health conditions including obesity, pre-existing diabetes, and chronic hypertension, derived from BORN records and validated ICES chronic disease cohorts. Maternal mental health and substance use history were captured using a previously validated composite measure, incorporating diagnoses of mood or anxiety disorders, psychotic disorders, substance use disorders, self-harm events, and related conditions identified through hospital admissions, emergency department visits, or multiple outpatient encounters in the two years preceding conception (38–41). Pregnancy-related characteristics included gestational diabetes, hypertensive disorders of pregnancy, mode of labour and delivery, and maternal and infant readmissions postpartum. Variables included in adjusted regression models were selected a priori based on clinical relevance and prior literature and are described below.

### Patient and Public Involvement

#### Statistical Analysis

Continuous variables were summarized using means and standard deviations, and categorical variables using counts and proportions. Outcome incidence rates per 1,000 pregnancies (95% confidence intervals) were calculated for each exposure group.

Relative risks and corresponding 95% confidence intervals were estimated using modified Poisson regression with robust error variance, an approach well suited for cohort studies with non-rare outcomes in which odds ratios may overestimate effect size. All adjusted models included covariates selected a priori based on clinical relevance and prior literature: maternal age, neighborhood income quintile, rurality, obesity, pre-existing diabetes and hypertension, parity, nicotine use, use of other substances, plurality, and fetal sex.

Analyses proceeded in three stages. First, we estimated associations between mutually exclusive opioid, OAT, and cannabis exposure groups and each outcome, using the no-substance-use group as the reference. Second, we assessed potential effect modification by fetal sex through inclusion of exposure-by–fetal sex interaction terms. Third, opioid, OAT, and cannabis exposures were modelled as separate predictors, and interactions between cannabis and opioid or OAT exposure were tested. Models were subsequently stratified by cannabis exposure to further assess effect modification.

Missing data were handled using complete-case analysis unless otherwise specified. Variables containing “unknown” categories followed prespecified recoding rules detailed in **Appendix 2**. Because some individuals contributed more than one pregnancy during the study period, models accounted for non-independence through clustering at the maternal level. All analyses were performed using SAS version 9.4 (Cary, North Carolina) at ICES Queen’s.

## Results

### Cohort Description

Between April 1^st^, 2013, to March 31^st^, 2021, there were 1,072,494 births in Ontario, Canada, of which 959,731 (89.5%) met eligibility criteria (**Figure 1**). Most included pregnancies (864,508; 90.1%) reported no substance exposure during pregnancy. Nicotine was the most common exposure (7.7%), followed by cannabis (2.4%), opioids (0.8%) and OAT (0.6 %). Cannabis co-exposure occurred in approximately 1 in 6 opioid- and/or OAT-exposed pregnancies.

**Figure 1.**
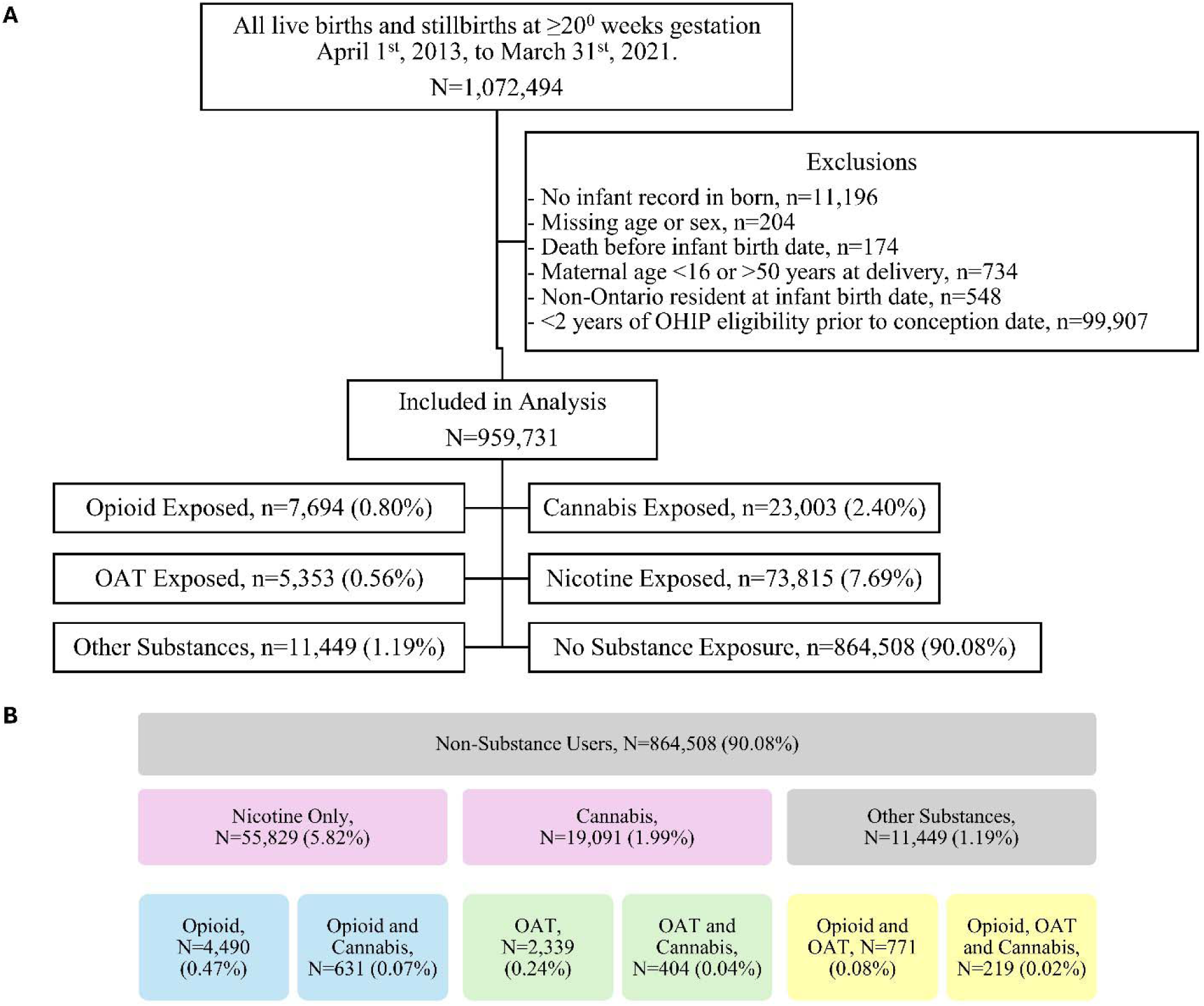
Cohort selection and exposure classification. A. Cohort derivation and overall exposure groups. Participants may appear in more than one exposure category; therefore, counts do not sum to the total sample size. **B.** Detailed, mutually exclusive exposure groups. Individuals in the opioid, OAT and cannabis exposure groups may also report nicotine use, and those in the other substance(s) may additionally report opioid, OAT, cannabis and/or nicotine use.

### Participant Characteristics

Maternal characteristics by exposure group are presented in **Table 1**. Compared to individuals with no substance exposure, those with substance exposure were generally younger, more often from lower income households, and more likely to live in a rural area. Participants reporting cannabis use were the youngest (mean age 26.4 ± 5.7 years). Parity distributions also differed across exposure groups; cannabis-exposed individuals had the highest proportion of primiparous pregnancies (51.5%). A history of mental illness was more common among all substance-exposed groups, ranging from 33.1% (nicotine) to 80.0% (OAT). A history of substance use disorder (within the past 2 years) was reported in 74.3% of OAT-exposed participants and 27.8% of opioid-exposed participants. Pregnancy complications varied by exposures, with the highest prevalence of gestational diabetes observed among those reporting “other substances” (9.6%) and the lowest proportion of placental abruption with no substance exposure (0.2%). Maternal readmission within 42 days of delivery was highest among opioid-exposed individuals (3.5%). Additional characteristics detailed by exposure groups can be found in **Supplemental Table 1**.

**Table 1.**
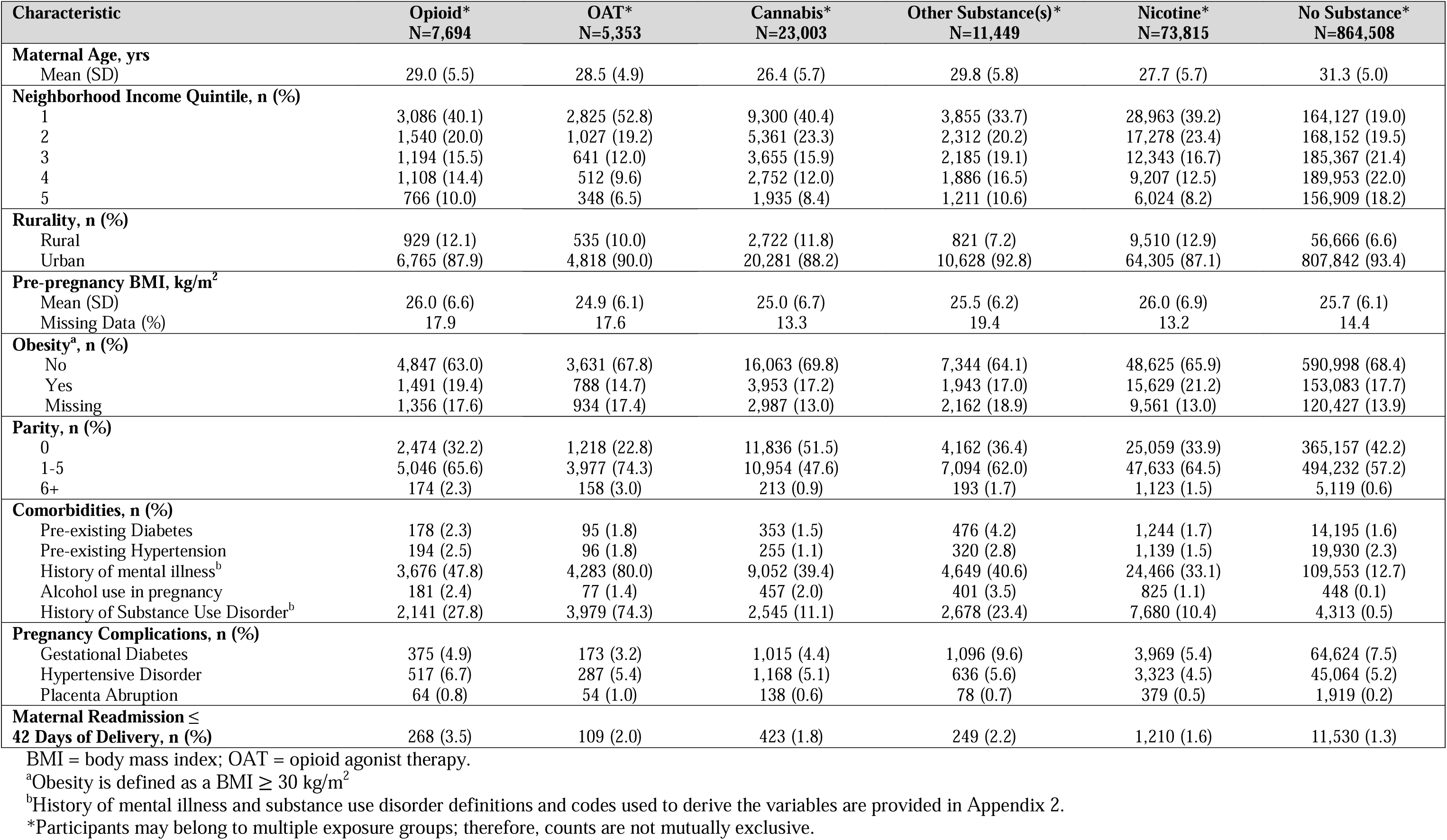
Maternal characteristics by substance exposure group.

### Fetal and Neonatal Characteristics

Fetal birth characteristics and outcomes by primary exposure group are shown in **Table 2**. Sex distribution and mean gestational age at delivery are similar across groups, whereas mean birthweight was highest in the no substance exposure group. All substance-exposed groups had higher proportions of SGA and IUGR compared with the no substance exposure group, with the highest proportions generally observed among those exposed to cannabis or OAT. SNM occurred in 7.2% of neonates in the no substance exposure group compared to 13.9% and 14.2% among opioid- and OAT-exposed neonates, respectively. NAS was diagnosed in 70.6% of OAT-exposed and 34.3% of opioid-exposed neonates. Jaundice and infant readmission within 42 days of delivery were most common among opioid- and OAT-exposed infants. Fetal and neonatal characteristics and health outcomes by detailed exposure groups are provided in **Supplemental Table 2**.

**Table 2.** Infant birth characteristics and neonatal outcomes by substance exposure group.

|  | Opioid* | OAT* | Cannabis* | Other<br>Substance(s)* | Nicotine* | No Substance * |
| --- | --- | --- | --- | --- | --- | --- |
|  | N=7,835 | N=5,412 | N=23,312 | N=11,644 | N=74,868 | N=879,090 |
| **Baby Sex, n (%)** |  |  |  |  |  |  |
| Female | 3,820 (48.58) | 2,637 (48.7) | 11,292 (48.4) | 5,711 (49.1) | 36,471 (48.7) | 428,242 (48.7) |
| Male | 4,015 (51.2) | 2,775 (51.3) | 12,027 (51.6) | 5,933 (50.9) | 38,397 (51.3) | 451,008 (51.3) |
| **Birthweight, g** |  |  |  |  |  |  |
| Mean (SD) | 3114.6 (686.6) | 3055.5 (662.2) | 3084.3 (652.6) | 3137.9 (646.6) | 3149.2 (618.4) | 3348.8 (591.6) |
| Female | 3061.6 (663.3) | 2985.2 (641.4) | 3033.7 (631.2) | 3083.5 (630.3) | 3087.7 (599.7) | 3289.5 (574.5) |
| Male | 3165.1 (704.4) | 3122.2 (674.7) | 3131.8 (668.6) | 3190.3 (657.7) | 3207.7 (630.0) | 3405.0 (601.9) |
| **Gestational Age, weeks** |  |  |  |  |  |  |
| Mean (SD) | 37.9 (2.6) | 37.9 (2.5) | 38.2 (2.5) | 38.1 (2.4) | 38.4 (2.3) | 38.7 (2.0) |
| **Small for Gestational Age, n (%)** |  |  |  |  |  |  |
| <3rd %ile | 364 (4.6) | 332 (6.1) | 1,433 (6.1) | 569 (4.9) | 3,985 (5.3) | 19,729 (2.2) |
| <10th %ile | 1,122 (14.3) | 983 (18.2) | 4,376 (18.8) | 1,779 (15.3) | 12,398 (16.6) | 79,082 (9.0) |
| **IUGR<sup>a</sup>, n (%)** | 300 (3.8) | 272 (5.0) | 1,255 (5.4) | 443 (3.8) | 3,087 (4.1) | 17,052 (1.9) |
| **Severe Neonatal Morbidity<sup>b</sup>, n (%)** | 1,087 (13.9) | 768 (14.2) | 2,656 (11.4) | 1,257 (10.8) | 6,867 (9.2) | 63,039 (7.2) |
| **Neonatal Abstinence Syndrome, n (%)** | 2,685 (34.3) | 3,823 (70.6) | 2,093 (9.0) | 2,493 (21.4) | 5,804 (7.8) | 1,012 (0.1) |
| **Jaundice, n (%)** | 1,470 (18.8) | 1,343 (24.8) | 3,075 (13.2) | 1,530 (13.1) | 8,174 (10.9) | 80,777 (9.2) |
| **Infant Readmission ≤**<br>**42 Days of Delivery, n (%)** | 797 (10.2) | 615 (11.4) | 1,878 (8.1) | 1,026 (8.8) | 5,234 (7.0) | 59,388 (6.8) |
| **Stillbirth, n (%)** | 38 (0.5) | 26 (0.5) | 85 (0.4) | 32 (0.3) | 251 (0.3) | 1,502 (0.2) |
| **Neonatal Mortality, n (%)** | 63 (0.8) | 51 (0.9) | 192 (0.8) | 70 (0.6) | 492 (0.7) | 3,633 (0.4) |

Descriptive data are presented at the infant level; therefore, denominators exceed the number of pregnancies because multiple gestations may contribute more than one infant.
OAT = opioid agonist therapy; IUGR = Intrauterine growth restriction
<sup>a</sup>IUGR was based on clinician diagnosis in BORN Ontario.
<sup>b</sup>Definitions of severe neonatal morbidity and code lists are provided in Appendix 2.
\*Participants may belong to multiple exposure groups; therefore, counts are not mutually exclusive.

### Primary Analyses

Incidence rates and relative risk of IUGR, SGA, and SNM by exposure groups are presented in **Table 3**. For both opioid- and OAT-exposed pregnancies, the incidence of SGA, IUGR, and SNM was consistently higher among those with cannabis co-exposure than among their non-cannabis-exposed counterparts. Most exposure groups had significantly increased adjusted risks across all outcomes relative to the no-substance-use group. Exceptions were the opioid-only, for which increased risk was limited to SNM, and the nicotine-only group, for which adjusted risks were not increased. For SGA outcomes, adjusted relative risk estimates were higher for opioid-plus-cannabis co-exposure group compared to opioid-only exposure, and similarly higher for OAT-plus-cannabis co-exposure compared with OAT-only exposure. There was no evidence of clinically meaningful effect modification by fetal sex across the primary outcomes (data not shown).

**Table 3.** Incidence and relative risk (RR) of fetal and neonatal outcomes by substance exposure group.

|  | Incidence per 1000 Pregnancies<br>(95% CI) | Unadjusted RR (95% CI) | Adjusted RR (95% CI)<sup>d</sup> |
| --- | --- | --- | --- |
| **Outcome: IUGR<sup>a</sup>** |  |  |  |
| **Opioid, OAT & Cannabis<sup>b</sup>** | **59.09 (31.46-101.05)** | **2.84 (1.68 - 4.81)** | **1.83 (1.07 - 3.15)** |
| Opioid & OAT<sup>b</sup> | 44.99 (31.34-62.57) | 2.12 (1.47 - 3.06) | 1.48 (1.03 - 2.13) |
| **Opioid & Cannabis<sup>b</sup>** | **54.43 (37.91-75.70)** | **2.82 (2.04 - 3.89)** | **1.77 (1.25 - 2.51)** |
| Opioid<sup>b</sup> | 26.17 (21.70-31.30) | 1.37 (1.15 - 1.64) | 1.14 (0.95 - 1.38) |
| **OAT & Cannabis<sup>b</sup>** | **51.34 (31.78-78.49)** | **2.66 (1.78 - 3.98)** | **1.70 (1.12 - 2.57)** |
| OAT<sup>b</sup> | 43.51 (35.52-52.77) | 2.31 (1.91 - 2.80) | 1.60 (1.30 - 1.97) |
| **Cannabis<sup>b</sup>** | **52.43 (49.25-55.76)** | **2.58 (2.41 - 2.75)** | **1.84 (1.68 - 2.01)** |
| Nicotine Only | 35.71 (34.17-37.30) | 1.80 (1.72 - 1.89) | 1.08 (0.97 - 1.20) |
| Other Substances<sup>c</sup> | 38.05 (34.58-41.76) | 1.96 (1.79 - 2.16) | 1.50 (1.34 - 1.66) |
| Non-Substance Users | 19.40 (19.11-19.69) | 1.00 (Ref.) | 1.00 (Ref.) |
| **Outcome: SGA <3%ile** |  |  |  |
| **Opioid, OAT & Cannabis<sup>b</sup>** | **72.73 (41.57-118.10)** | **3.22 (2.00 - 5.17)** | **1.73 (1.06 - 2.81)** |
| Opioid & OAT<sup>b</sup> | 64.27 (47.70-84.73) | 2.82 (2.15 - 3.70) | 1.62 (1.22 - 2.16) |
| **Opioid & Cannabis<sup>b</sup>** | **74.65 (55.04-98.98)** | **3.28 (2.49 - 4.33)** | **1.84 (1.37 - 2.47)** |
| Opioid<sup>b</sup> | 28.14 (23.49-33.43) | 1.27 (1.07 - 1.50) | 1.01 (0.85 - 1.19) |
| **OAT & Cannabis<sup>b</sup>** | **85.57 (59.61-119.01)** | **3.70 (2.67 - 5.14)** | **2.07 (1.47 - 2.91)** |
| OAT<sup>b</sup> | 47.74 (39.34-57.40) | 2.13 (1.77 - 2.56) | 1.36 (1.11 - 1.65) |
| **Cannabis<sup>b</sup>** | **58.48 (55.12-61.99)** | **2.54 (2.39 - 2.70)** | **1.61 (1.48 - 1.75)** |
| Nicotine Only | 46.90 (45.14-48.72) | 2.06 (1.98 - 2.15) | 1.07 (0.97 - 1.18) |
| Other Substances<sup>c</sup> | 48.87 (44.93-53.05) | 2.14 (1.97 - 2.33) | 1.50 (1.36 - 1.66) |
| Non-Substance Users | 22.44 (22.13-22.76) | 1.00 (Ref) | 1.00 (Ref) |
| **Outcome: SGA <10%ile** |  |  |  |
| **Opioid, OAT & Cannabis<sup>b</sup>** | **231.82 (172.60-304.80)** | **2.50 (1.96 - 3.19)** | **1.63 (1.26 - 2.10)** |
| Opioid & OAT<sup>b</sup> | 186.38 (157.27-219.30) | 2.02 (1.74 - 2.34) | 1.39 (1.19 - 1.62) |
| **Opioid & Cannabis<sup>b</sup>** | **199.07 (166.08-236.69)** | **2.13 (1.81 - 2.49)** | **1.43 (1.21 - 1.69)** |
| Opioid<sup>b</sup> | 107.09 (97.82-116.99) | 1.21 (1.11 - 1.31) | 1.05 (0.97 - 1.14) |
| **OAT & Cannabis<sup>b</sup>** | **239.61 (194.53-292.01)** | **2.55 (2.13 - 3.06)** | **1.72 (1.43 - 2.07)** |
| OAT<sup>b</sup> | 152.94 (137.59-169.53) | 1.71 (1.55 - 1.88) | 1.27 (1.15 - 1.41) |
| **Cannabis<sup>b</sup>** | **183.19 (177.20-189.32)** | **1.96 (1.90 - 2.02)** | **1.44 (1.38 - 1.50)** |
| Nicotine Only | 150.28 (147.10-153.51) | 1.64 (1.60 - 1.67) | 1.03 (0.98 - 1.08) |
| Other Substances<sup>c</sup> | 152.78 (145.76-160.05) | 1.66 (1.59 - 1.74) | 1.32 (1.25 - 1.39) |
| Non-Substance Users | 89.96 (89.33-90.59) | 1.00 (Ref) | 1.00 (Ref) |
| **Outcome: SNM** |  |  |  |
| **Opioid, OAT & Cannabis<sup>b</sup>** | **154.55 (107.03-215.96)** | **2.15 (1.55 - 2.99)** | **1.92 (1.40 - 2.63)** |
| Opioid & OAT<sup>b</sup> | 134.96 (110.39-163.38) | 1.84 (1.52 - 2.22) | 1.62 (1.34 - 1.96) |
| **Opioid & Cannabis<sup>b</sup>** | **169.52 (139.19-204.49)** | **2.39 (2.01 - 2.85)** | **1.91 (1.60 - 2.28)** |
| Opioid<sup>b</sup> | 121.70 (111.81-132.23) | 1.64 (1.51 - 1.78) | 1.51 (1.38 - 1.64) |
| **OAT & Cannabis<sup>b</sup>** | **156.48 (120.51-199.82)** | **2.10 (1.66 - 2.67)** | **1.93 (1.52 - 2.45)** |
| OAT<sup>b</sup> | 125.90 (112.01-141.03) | 1.74 (1.56 - 1.95) | 1.62 (1.43 - 1.82) |
| **Cannabis<sup>b</sup>** | **106.25 (101.71-110.95)** | **1.46 (1.40 - 1.53)** | **1.26 (1.19 - 1.33)** |
| Nicotine Only | 80.65 (78.33-83.02) | 1.12 (1.08 - 1.15) | 0.89 (0.84 - 0.95) |
| Other Substances<sup>c</sup> | 107.95 (102.07-114.09) | 1.47 (1.39 - 1.56) | 1.32 (1.24 - 1.40) |
| Non-Substance Users | 71.71 (71.15-72.27) | 1.00 (Ref) | 1.00 (Ref) |

OAT = opioid agonist therapy; IUGR = Intrauterine growth restriction; SGA = small for gestational age; SNM = severe neonatal morbidity
<sup>a</sup> IUGR was based on clinician diagnosis in BORN Ontario.
<sup>b</sup> Participants in the opioid, OAT, cannabis, and combination groups may also have nicotine exposure; groups are mutually exclusive with respect to opioid, OAT, and cannabis exposure.
<sup>c</sup> The “Other Substances” group may include concurrent opioid, OAT, cannabis, and/or nicotine exposure.
<sup>d</sup> Model adjusted for maternal age, income, rurality, obesity, pre-existing diabetes and hypertension, parity, nicotine exposure, other substance use, singleton or multiple, and fetal sex.

### Individual Exposure and Interaction Analyses

When opioid, OAT, and cannabis exposures were examined independently in the models (**Supplemental Table 3**), cannabis exposure was associated with significantly increased risk across all outcomes, with adjusted relative risks ranging from a 35% increased risk for SNM (aRR=1.35, 95% CI 1.30-1.41) to a 77% increased risk for IUGR (aRR=1.77, 95% CI 1.66-1.89). OAT exposure was also associated with elevated risks, ranging from a 23% increased risk for SGA <10^th^ percentile (aRR=1.23, 95% CI 1.14-1.32) to 58% increased risk of SNM (aRR=1.58, 95% CI 1.45-1.73). For opioid exposure, significantly increased risk was observed only for SNM (aRR=1.51, 95% CI 1.41-1.62). Significant interaction terms were identified for opioid*cannabis (SGA <10^th^ percentile and SNM) and OAT*cannabis (IUGR). However, in cannabis-stratified models (**Supplemental Table 4**), adjusted risk estimates were similar across strata, suggesting limited evidence of effect modification by cannabis exposure.

## Discussion

### Statement of principal findings

In this large population-based cohort, pregnancies exposed to opioids, OAT, and cannabis had increased rates of adverse fetal and neonatal outcomes, although the magnitude and pattern of risk differed by exposure. Cannabis co-exposure was consistently associated with higher incidences of IUGR, SGA (<3^rd^ and <10^th^ percentile), and SNM among pregnancies exposed to opioids or OAT compared with their corresponding non-cannabis-exposed groups.

When exposures were considered separately, opioid exposure without OAT or cannabis co-exposure was not significantly associated with IUGR or SGA after adjustment, although the risk of SNM remained elevated. OAT exposure was associated with modestly increased risks of fetal growth outcomes and SNM. Cannabis exposure without opioid of OAT co-exposures was associated with increased risks across all fetal growth outcomes and SNM, with effect estimates for fetal growth similar to or greater than those observed with opioid or OAT exposure alone. These findings highlight both the importance of accounting for co-exposures when evaluating opioid-related pregnancy outcomes and the substantial fetal growth risks associated with prenatal cannabis exposure.

### Strengths and limitations of the study

This study has several notable strengths. First, it leverages one of the largest population-based evaluations of opioid, OAT, and cannabis exposure in pregnancy, drawing on a validated provincial perinatal registry capturing more than 99% of hospital births in Ontario (42, 43). Second, linkage of BORN Ontario with multiple administrative health databases at ICES enabled comprehensive ascertainment of maternal health histories and fetal and neonatal outcomes while minimizing loss to follow-up. Third, the large sample size permitted examination of clinically relevant, mutually exclusive opioid, OAT, and cannabis exposure combinations that have been difficult to evaluate in previous studies. Finally, adjusted analyses accounted for several important potential confounders, including maternal age, socioeconomic status, rurality, obesity, pre-existing diabetes and hypertension, parity, nicotine and other substance exposure, plurality, and fetal sex.

Several limitations should also be considered. Substance exposure was primarily identified through maternal self-report and provider documentation and is therefore likely to underestimate true exposure. Such misclassification may have resulted in some exposed pregnancies being classified as unexposed and would generally be expected to attenuate observed associations. Detailed information regarding substance dose, frequency, route of administration, potency, and timing across gestation was unavailable, limiting assessment of dose-response and trimester-specific effects. Although analyses adjusted for multiple clinical and sociodemographic factors, residual confounding from unmeasured or incompletely measured characteristics - including psychosocial factors and patterns of polysubstance use - cannot be excluded. Additionally, SGA was defined using population-based birthweight references that do not account for individual constitutional or race- or ethnicity-related variation in fetal growth and may therefore result in some misclassification (44). IUGR was based on clinician documentation in BORN Ontario and, as such, may reflect variation in clinical recognition and diagnostic practice rather than a standardized research definition. OAT exposure was captured as a single category and did not permit reliable differentiation of methadone- and buprenorphine-based regimens; this is important because fetal and neonatal outcomes may differ between these treatments, with several comparative studies and meta-analyses reporting more favorable growth and neonatal withdrawal outcomes following buprenorphine exposure than methadone exposure (26, 29). Finally, as with all observational studies, the associations identified cannot establish causality.

### Strengths and weaknesses in relation to other studies, discussing important differences in results

Prior research evaluating opioid exposure during pregnancy has consistently demonstrated associations with adverse perinatal outcomes, including impaired fetal growth, preterm birth, neonatal opioid withdrawal, and other neonatal morbidity (45, 46). OAT requires a distinct interpretation. Although pregnancies treated with OAT remain at increased risk for outcomes such as impaired fetal growth and neonatal withdrawal relative to pregnancies without opioid exposure, methadone and buprenorphine remain the recommended treatments for opioid use disorder during pregnancy. Maintenance therapy stabilizes maternal opioid exposure and reduces the risks associated with ongoing non-prescribed opioid use, recurrent withdrawal, and relapse; medically supervised withdrawal is generally not recommended because of high relapse rates and associated maternal and fetal risks (25, 47). Accordingly, the elevated risks observed among OAT-exposed pregnancies in this study should not be interpreted as evidence that OAT itself is more harmful than untreated opioid use. Rather, they likely reflect a combination of medication exposure, the underlying opioid use disorder for which OAT is prescribed, associated comorbidities and social determinants of health, and concurrent substance exposures.

An important contribution of the present study is the ability to examine cannabis co-exposure within opioid- and OAT-exposed pregnancies. Multi-substance use is common among individuals with opioid-use disorder, yet much of the existing literature has examined opioid or OAT exposure without adequately characterizing concurrent cannabis use. Smaller cohorts, including ENRICH-1 (n=251), have demonstrated a high prevalence of cannabis use among pregnant individuals with opioid use disorder (43.2%), but were not designed or powered to determine the independent contribution of cannabis co-exposure to fetal-growth outcomes (48). Studies of specialized perinatal substance-use programs have similarly demonstrated that cannabis use is common among individuals receiving treatment for opioid use disorder and may vary according to treatment and patterns of opioid use (49). A retrospective study by Stein et al. reported greater odds of low birth weight among pregnancies with concurrent opioid and cannabis exposure than among those with opioid exposure alone (25.7% vs 19.1%; aOR 1.46, [1.1-1.9]), although interpretation was limited by small sample size and substantial psychiatric comorbidity (50). Collectively, the literature indicates that co-exposure is common and clinically important, yet the magnitude of its contribution to fetal growth restriction remains insufficiently characterized.

Our findings extend this literature in a large population-based cohort. Among pregnancies exposed to opioids or OAT, those with cannabis co-exposure generally had higher incidences of IUGR, SGA, and SNM than their corresponding non-cannabis-exposed groups. However, the formal interaction and cannabis-stratified analyses did not provide consistent evidence that cannabis biologically potentiates the effects of opioids or OAT. Thus, cannabis co-exposure appears to identify a particularly vulnerable clinical population with a greater burden of adverse outcomes, but these data do not establish a synergistic interaction between the substances. This distinction is clinically important: concurrent cannabis use should be recognized as a marker of additional fetal and neonatal risk without attributing that increased risk solely to a pharmacological interaction.

The study also highlights the risks associated with prenatal cannabis exposure independent of opioids or OAT. Although earlier reviews reported heterogeneous findings and raised concerns regarding residual confounding, particularly by tobacco use, more recent systematic reviews and large population-based studies have provided increasingly consistent evidence of impaired fetal growth associated with prenatal cannabis exposure (32, 33, 51–53). A 2022 systematic review and meta-analysis, for example, reported approximately twice the risk of low birthweight (< 2500g; RR 2.06, 95%CI 1.25 - 3.42), a 61% greater risk of SGA (≤5%ile; RR 1.61, 95% CI 1.44-1.79), and a mean reduction in birthweight of 112g (95% CI, -167 to -57g) among cannabis-exposed pregnancies (54). More recently, a 2025 meta-analysis of adjusted estimates from over 21 million pregnancies reported increased odds of both low birthweight (aOR 1.75, 95% CI 1.41–2.18) and SGA (aOR 1.57, 95% CI 1.36–1.81), with the certainty of evidence for these associations now rated as moderate (32). Large Canadian population-based studies have similarly demonstrated higher rates of low birthweight and SGA among cannabis-exposed pregnancies after adjustment for other substance use and relevant maternal characteristics (33, 55).

Remaining variability across studies may reflect differences in exposure ascertainment, timing and frequency of use, underreporting, residual confounding, co-exposures, and changes in cannabis potency over time. THC concentrations in cannabis products have increased substantially over recent decades, raising the possibility that contemporary prenatal exposures may not be directly comparable with those captured in older cohorts (56). Although temporal changes in fetal growth effects have not, to our knowledge, been formally evaluated in relation to changing THC potency, a recent prospective study quantifying exposure to contemporary high-potency cannabis products reported lower gestational age-adjusted birthweight and length, with larger differences among individuals who continued frequent cannabis use throughout pregnancy (57).

Our findings extend this evolving literature by demonstrating substantial associations between cannabis exposure and fetal-growth outcomes within a population specifically enriched for opioid and OAT exposure. In the independent-exposure models, cannabis was associated with a 77% increased adjusted risk of IUGR, a 54% increased risk of SGA <3rd percentile, a 39% increased risk of SGA <10th percentile, and a 35% increased risk of SNM. These associations were comparable to, and for several fetal-growth outcomes greater than, those observed for opioid or OAT exposure. Luke et al. reported evidence of effect modification by infant sex for SGA, with greater susceptibility among cannabis-exposed female infants (33); in contrast, we found no clear evidence that the associations in our cohort differed meaningfully by fetal sex. Together, these findings reinforce that cannabis should not be considered a benign exposure during pregnancy and demonstrate that its association with impaired fetal growth is evident both independently and in pregnancies complicated by opioid use or OAT.

### Meaning of the study: possible explanations and implications for clinicians and policymakers

These findings have several implications for clinical care. First, assessment of substance use during pregnancy should extend beyond identification of opioid use or OAT to include cannabis, nicotine, and other concurrent substances. This is particularly relevant in jurisdictions such as Ontario, where recreational cannabis is legal and readily accessible. Legalization may contribute to normalization of cannabis use and lower perceived risk, and pregnant individuals may use cannabis to manage pregnancy-related symptoms including nausea, anxiety, sleep disturbance, and pain (58–60). In one survey, more than half of pregnant individuals who continued to use cannabis considered its use during pregnancy to be safe, while alcohol and tobacco were more consistently perceived as harmful (60). Canadian studies conducted before and after recreational cannabis legalization have also examined changes in prenatal cannabis use following legalization, underscoring the importance of considering the evolving legal and social context in which cannabis exposure occurs (58). At the same time, contemporary reproductive-age populations have access to cannabis products with substantially higher THC concentrations than those available in earlier decades (56), meaning that the exposure landscape captured by older pregnancy studies may not fully reflect current use. The long-term developmental consequences of these contemporary prenatal exposures remain incompletely characterized. In this context, the present findings provide clinically relevant evidence for counselling that cannabis should not be considered a benign exposure during pregnancy and that prenatal cannabis exposure is associated with measurable risks to fetal growth, including when used without concurrent opioid or OAT exposure.

Second, the findings should not discourage the use of OAT in pregnancy. OAT remains the standard of care for opioid use disorder and an essential component of harm reduction. Rather, individuals receiving OAT may benefit from comprehensive, non-stigmatizing assessment of concurrent substance use and the circumstances contributing to it. Where cannabis co-use is identified, counselling can address the observed associations with impaired fetal growth and neonatal morbidity while exploring the reasons for use and supporting reduction or cessation when feasible. Abrupt opioid withdrawal should not be promoted as a strategy to reduce fetal exposure.

Finally, the marked differences in maternal characteristics across exposure groups emphasize the importance of addressing substance use within its broader clinical and social context. Mental illness and substance use disorder were substantially more prevalent among substance-exposed participants than among those reporting no substance exposure, and socioeconomic disadvantage was also more common in several exposure groups. Integrated prenatal care that combines obstetric care, addiction treatment, mental-health services, and appropriate social supports may therefore be particularly important for this population. The objective should be not simply to identify individual substance exposures, but to provide care that addresses the constellation of factors associated with maternal and infant risk.

### Unanswered questions and future research

Several questions remain. The biological mechanisms linking opioids, OAT, and cannabis with impaired fetal growth are incompletely understood. The placenta represents one potential mediator because both opioid and cannabinoid signaling pathways can influence placental development and function, including trophoblast signaling, vascularization, nutrient exchange, and endocrine activity; however, the relative contributions of direct fetal exposure and placenta-mediated effects remain incompletely understood (61, 62).

Future research should prioritize prospective and repeated measurement of substance exposure across pregnancy, including dose, frequency, potency, route of administration, indication for use, and timing of initiation, continuation, reduction, or cessation. Such data would help distinguish persistent from intermittent exposure and permit assessment of dose-response and gestational timing effects that could not be examined in the present study. More detailed characterization of OAT is also needed, including differentiation between methadone and buprenorphine and consideration of treatment stability and concurrent non-prescribed opioid use. Further work is also needed to understand why cannabis co-exposure identifies pregnancies with particularly high rates of adverse outcomes. Larger prospective studies incorporating measures of mental health, nutrition, housing and socioeconomic circumstances, treatment engagement, and other substance use may help determine the extent to which the observed risk reflects cannabis exposure itself versus the broader clinical and social context in which co-use occurs. Longitudinal follow-up of exposed children will additionally be important to determine whether the fetal-growth differences observed in this study translate into later neurodevelopmental, metabolic, or cardiovascular consequences, particularly given emerging but heterogeneous evidence of longer-term effects following prenatal opioid and cannabis exposure and the established links between impaired fetal growth and later-life cardiometabolic risk (63–65).

### Conclusion

In this large population-based cohort, pregnancies exposed to opioids, OAT, and cannabis had increased risks of impaired fetal growth and severe neonatal morbidity, with the highest observed risks generally occurring among opioid- or OAT-exposed pregnancies with concurrent cannabis exposure. However, formal interaction and stratified analyses provided limited evidence that cannabis modified the effects of opioid or OAT exposure, suggesting that co-exposure identifies a particularly high-risk population rather than establishing a synergistic effect. Cannabis exposure was also independently associated with IUGR, SGA, and SNM, with fetal-growth risk estimates comparable to or greater than those observed with opioid or OAT exposure. These findings support comprehensive assessment of concurrent substance use during pregnancy, continued use of evidence-based OAT for opioid use disorder, and non-stigmatizing counselling regarding the fetal and neonatal risks associated with cannabis exposure.

## Ethical Statement

This study was reviewed for ethical compliance by the Queen’s University Health Sciences and Affiliated Teaching Hospitals Research Ethics Board and received initial clearance on February 8^th^, 2024 (Reference number# 6040679).

## Transparency Statement

The corresponding author affirms that this manuscript is an honest, accurate, and transparent account of the study being reported; that no important aspects of the study have been omitted; and that any discrepancies from the study as planned (and, if relevant, registered) have been explained.

## Statement of Authorship

All authors have made substantial contributions to the conception, design of the work, the acquisition of data and the interpretation of results. All authors have drafted the work or revised it critically for important intellectual content. All authors have approved the final version submitted for publication and all agree to be accountable for all aspects of the work in ensuring that questions related to the accuracy or integrity of any part of the work are appropriately investigated and resolved.

## Acknowledgements

Parts of this material are based on data and/or information compiled and provided by CIHI and the Ontario Ministry of Health. The analyses, conclusions, opinions and statements expressed herein are solely those of the authors and do not reflect those of the funding or data sources; no endorsement is intended or should be inferred.

Parts of this material are based on data and information provided by Ontario Health (OH). The opinions, results, view, and conclusions reported in this paper are those of the authors and do not necessarily reflect those of OH. No endorsement by OH is intended or should be inferred.

This document used data adapted from the Statistics Canada Postal Code^OM^ Conversion File, which is based on data licensed from Canada Post Corporation, and/or data adapted from the Ontario Ministry of Health Postal Code Conversion File, which contains data copied under license from ©Canada Post Corporation and Statistics Canada.

## Data availability statement

The dataset from this study is held securely in coded form at ICES. Although data-sharing agreements prohibit ICES from making the dataset publicly available, access may be granted to those who meet prespecified criteria for confidential access, available at www.ices.on.ca/DAS. The full dataset creation plan and underlying analytic code are available from the authors upon request, understanding that the computer programmes may rely upon coding templates or macros that are unique to ICES and therefore either inaccessible or requiring modification.

