## Appendix and Supplement for "The Impact of Opioid, Opioid Agonist Therapy, and Cannabis Exposure on Fetal Growth: A Population-Based Cohort Study"

**Appendix 1** – Overview of administrative and clinical data sources used in the study.

| **Dataset** | **Description** | **Key Contribution to the Study** |
|---|---|---|
| **ICES Data Environment** | | |
| | ICES houses linked, population-based administrative health datasets for Ontario residents. Using encrypted identifiers, data from BORN Ontario were linked with hospital, emergency, outpatient, demographic, and chronic disease datasets to identify exposures, maternal histories, and fetal and neonatal outcomes. | This environment enabled the integration of perinatal records with maternal substance exposure, health histories, demographic information, and maternal–infant outcomes for this study. |
| **Perinatal Dataset** | | |
| Better Outcomes Registry & Network (BORN) Ontario | A longitudinal administrative data source that collects information related to maternal, perinatal, and newborn health in Ontario. Includes a six-year dataset (2006-2011) of legacy birth record data (formerly collected in the Niday Perinatal Database). | Primary source of perinatal data, including substance exposures, maternal characteristics, pregnancy complications, fetal growth measures, and neonatal outcomes. |
| **Administrative Health Databases** | | |
| Canadian Institute for Health Information Discharge Abstract Database (CIHI DAD) | A dataset that captures administrative, clinical, and demographic information on hospital discharges, including deaths, sign-outs, and transfers. | Used to identify maternal hospitalizations, comorbidities, and maternal/infant readmissions. |
| National Ambulatory Care Reporting System (NACRS) | A dataset that contains data on hospital-based and community-based ambulatory care: day surgery, outpatient and community-based clinics, and emergency departments. | Used to capture emergency department visits and outpatient procedures relevant to mental health and substance use. |
| Ontario Health Insurance Plan Claims Database (OHIP) | A dataset that records all claims for reimbursement by Ontario physicians for inpatient and ambulatory visits, consultations and procedures. The data also include claims from optometrists for publicly funded reimbursement and | Used to identify outpatient encounters and physician-billed diagnoses. |

| Dataset | Description | Key Contribution to the Study |
|---|---|---|
| | from laboratories for all diagnostic tests performed. | |
| Ontario Mental Health Reporting System (OMHRS) | The Ontario Mental Health Reporting System (OMHRS) in Ontario officially collects data on patients in adult designated inpatient mental health beds. This includes beds in General, Provincial Psychiatric, and Specialty Psychiatric facilities. | Provided inpatient mental-health admissions used to identify maternal mental illness and substance use disorder history. |
| **Population and Demographic Databases** | | |
| Registered Persons Database (RPDB) | A dataset that provides demographic information about all individuals who have received an Ontario health card number, including their date of birth, sex, and home address. | Used for demographic linkage, residential postal code, and mortality data. |
| Postal Code Conversion File (PCCF) | A digital file which provides a correspondence between the Canada Post Corporation (CPC) six-character postal code and Statistics Canada's standard geographic areas for which census data and other statistics are produced. | Used to map residential postal codes to geographic areas, enabling assignment of neighborhood income quintile and rurality. |
| **Derived Cohorts** | | |
| Ontario Hypertension Dataset (HYPER) | A dataset of all of the people in Ontario identified as having hypertension. | Provided validated records of pre-existing hypertension used to classify maternal chronic hypertension prior to pregnancy. |
| Linked Delivering Mothers and Newborns (MOMBABY) | A dataset that is derived within ICES to link the inpatient admission records of delivering mothers and their newborns. | Enabled linkage of maternal and newborn hospital records and contributed additional diagnoses and procedures used to ascertain neonatal outcomes, including SNM and NAS. |
| Ontario Diabetes Dataset (ODD) | A dataset of all of the people in Ontario diagnosed with diabetes. | Provided validated records of pre-existing diabetes used to classify maternal chronic |

| Dataset | Description | Key Contribution to the Study |
|---|---|---|
| | | diabetes prior to pregnancy. |

**Appendix 2** – Definitions of exposures, outcomes, and covariates, including diagnostic and procedural code lists.

| | Description | Applicable codes | Data Sources |
|---|---|---|---|
| **Cohort Cut** | | | |
| **Inclusion criterion** | All livebirths and stillbirths at ≥$20^0$ weeks gestation among women aged 16-50 years. April 1st, 2013, to March 31st, 2021. | -- | BORN |
| **Exclusion Criteria** | No IKN or inability to link to ICES datasets. | -- | RPDB |
| | Birth outside of Ontario. | -- | BORN |
| | No infant record in BORN. | -- | BORN |
| | Less than 2 years of OHIP eligibility prior to the estimated date of conception. | = estimated_bdate – (40*7) | RPDB |
| **Exposures** | | | |
| **Opioid Exposure** | Yes<br><br>*If one variable = YES and the other variable = UNKNOWN, code the exposure as YES* | EXPOS_DRUG_AND_SUBST_ID=<br>1020520 (opioids)<br>1020210 (Narcotics, prior to April 2014)<br>**OR**<br>EXPOSURE_MEDICATION_ID=<br>1020700 = Opioids | BORN |
| | No | EXPOS_DRUG_AND_SUBST_ID≠<br>1020520 (opioids)<br>1020210 (Narcotics, prior to April 2014)<br>1020535 = Unknown<br>**AND**<br>EXPOSURE_MEDICATION_ID≠<br>1020700 = Opioids<br>1020795 = Unknown | BORN |
| | Unknown | EXPOS_DRUG_AND_SUBST_ID= | BORN |

| | Description | Applicable codes | Data Sources |
|---|---|---|---|
| | *For analysis, group Unknown with No* | 1020535 = Unknown<br>**AND**<br>EXPOSURE_MEDICATION_ID=<br>1020795 = Unknown | |
| **Opioid agonist therapy (OAT) Exposure** | Yes<br><br>*If one variable = YES and the other variable = UNKNOWN, code the exposure as YES* | EXPOS_DRUG_AND_SUBST_ID=<br>1020500 = Methadone (prior to April 2014)<br>**OR**<br>EXPOSURE_MEDICATION_ID=<br>1020715 = Methadone/Subutex | BORN |
| | No | EXPOS_DRUG_AND_SUBST_ID≠<br>1020500 = Methadone (prior to April 2014)<br>1020535 = Unknown<br>**AND**<br>EXPOSURE_MEDICATION_ID≠<br>1020715 = Methadone/Subutex<br>1020795 = Unknown | BORN |
| | Unknown<br><br>*For analysis, group Unknown with No* | EXPOS_DRUG_AND_SUBST_ID=<br>1020535 = Unknown<br>**AND**<br>EXPOSURE_MEDICATION_ID=<br>1020795 = Unknown | BORN |
| **Cannabis Exposure** | Yes<br><br>*If one variable = YES and the other variable = UNKNOWN, code the exposure as YES* | EXPOS_DRUG_AND_SUBST_ID=<br>1020490 = Marijuana (prior to April 2019)<br>**OR**<br>CANNABIS_EXPOSURE_FLAG= YES | BORN |
| | No | EXPOS_DRUG_AND_SUBST_ID≠<br>1020490 = Marijuana (prior to April 2019)<br>**AND**<br>CANNABIS_EXPOSURE_FLAG= NO | BORN |
| | Unknown<br><br>*For analysis, group Unknown with No* | EXPOS_DRUG_AND_SUBST_ID=<br>1020535 = Unknown<br>**AND**<br>CANNABIS_EXPOSURE_FLAG=<br>UNKNOWN | BORN |
| **Nicotine Exposure** | Yes | MAT_SMOKING_AT_ADM_FOR_BIRTH_ID=<br>1017390 = <10 cigarettes/day | BORN |

| | Description | Applicable codes | Data Sources |
|---|---|---|---|
| | | 1017400 = 10-20 cigarettes/day<br>1017410 = >20 cigarettes/day<br>1017420 = Amount unknown | |
| | No | MAT_SMOKING_AT_ADM_FOR_BIRTH_ID= 1017380 = None | BORN |
| | Unknown<br><br>*For analysis, group Unknown with No* | MAT_SMOKING_AT_ADM_FOR_BIRTH_ID= 1017425 = Unknown | BORN |
| **Other Substance Exposure** | Yes<br><br>*If one variable = YES and the other variable = UNKNOWN, code the exposure as YES* | EXPOS_DRUG_AND_SUBST_ID=<br>1020460 = Cocaine<br>1020470 = Gas/Glue<br>1020480 = Hallucinogens<br>1020530 = Other<br>**OR**<br>EXPOSURE_MEDICATION_ID=<br>1020540 = Amphetamines | BORN |
| | No | EXPOS_DRUG_AND_SUBST_ID≠<br>1020460 = Cocaine<br>1020470 = Gas/Glue<br>1020480 = Hallucinogens<br>1020530 = Other<br>**AND**<br>EXPOSURE_MEDICATION_ID≠<br>1020540 = Amphetamines | BORN |
| | Unknown<br><br>*For analysis, group Unknown with No* | EXPOS_DRUG_AND_SUBST_ID=<br>1020535 = Unknown<br>**AND**<br>EXPOSURE_MEDICATION_ID=<br>1020795 = Unknown | BORN |
| **No Substance Use Exposure*** | Yes | EXPOS_DRUG_AND_SUBST_ID=<br>1020457 = None<br>**AND**<br>EXPOSURE_MEDICATION_ID≠<br>1020700 = Opioids, 1020715 = Methadone/Subutex, 1020540 = Amphetamines<br>**AND**<br>CANNABIS_EXPOSURE_FLAG= NO<br>**AND**<br>MAT_SMOKING_AT_ADM_FOR_BIRTH_ID=<br>1017380 = None | BORN |
| | No | EXPOS_DRUG_AND_SUBST_ID≠ | BORN |

| | Description | Applicable codes | Data Sources |
|---|---|---|---|
| | | 1020457 = None<br>**OR**<br>EXPOSURE_MEDICATION_ID=<br>1020700 = Opioids, 1020715 = Methadone/Subutex, 1020540 = Amphetamines<br>**OR**<br>CANNABIS_EXPOSURE_FLAG=YES<br>**OR**<br>MAT_SMOKING_AT_ADM_FOR_BIRTH_ID=<br>1017390 = <10 cigarettes/day<br>1017400 = 10-20 cigarettes/day<br>1017410 = >20 cigarettes/day<br>1017420 = Amount unknown | |
| | Unknown<br><br>*For analysis, group Unknown with Yes* | EXPOS_DRUG_AND_SUBST_ID=<br>1020535 = Unknown<br>**AND**<br>EXPOSURE_MEDICATION_ID=<br>1020795 = Unknown<br>**AND**<br>CANNABIS_EXPOSURE_FLAG=<br>UNKNOWN<br>**AND**<br>MAT_SMOKING_AT_ADM_FOR_BIRTH_ID=<br>1017425 = Unknown | BORN |
| **Outcome Measures**<br>*Outcomes were modelled per pregnancy, if at least one baby in a set of multiples experienced an outcome the pregnancy was considered positive for that outcome.* | | | |
| **Primary Outcomes** | | | |
| **Small for Gestational Age** | <3rd and <10th percentile for sex and gestational age.<br><br>*If birthweight, sex or gestational age are missing, exclude from outcome.* | Kramer MS, Demissie K, Yang H, Platt RW, Sauvé R, Liston R. The contribution of mild and moderate preterm birth to infant mortality. Fetal and Infant Health Study Group of the Canadian Perinatal Surveillance System. JAMA: the journal of the American Medical Association. 2000;284(7):843-9. | BORN |
| **Intrauterine Growth Restriction** | Yes/No | NEONATAL_HEALTH_CONDITIONS_ID | BORN |

| | Description | Applicable codes | Data Sources |
|---|---|---|---|
| | | = 1026966 = Other Health Conditions \ Intrauterine Growth Restriction (IUGR)<br>OR<br>COMPLICATION_ID<br>= 1020210 = Fetal \ IUGR | |
| **Secondary Outcomes** | | | |
| **Stillbirth** | Yes/No | Stillbirth<br>(AGG_PREGNANCY_OUTCOME_ID)<br>Any of the following:<br>1021060 = Stillbirth at >=20wks or >=500gms<br>1021070 = Stillbirth at >=20wks or >=500gms \ Termination<br>1021080 = Stillbirth at >=20wks or >=500gms \ Spontaneous - Occurred during antepartum period<br>1021090 = Stillbirth at >=20wks or >=500gms \ Spontaneous - Occurred during intrapartum period | BORN |
| **Neonatal Mortality** | Death within 28 days of delivery | In BORN: NEONATAL_DEATH_ID = 1018200 = Yes<br>In RPDB: DTH=Yes and MOHDATE within 28 days of delivery | BORN and RPDB |
| **Neonatal Abstinence Syndrome** | Yes/No | In BORN: NEONATAL_HEALTH_CONDITIONS_ID = 1026410<br>In MOMBABY: P96.1 (Neonatal withdrawal symptoms from maternal use of drugs of addiction | BORN and MOMBABY |
| **Severe Neonatal Morbidity** | Based on use of ICD-10 and CCI codes to capture neonatal morbidity components during birth admission and up to 28 days of life. | • Gestational age <32 weeks<br>• Birthweight <1,500 g<br>**ICD-10-CA Diagnosis Code**<br>• Birth trauma (intracranial hemorrhage paralysis due to brachial plexus injury, skull or long bone fracture): P100 TO P103, P130, P132, P133, P140, P141<br>• Broncho-pulmonary dysplasia: P271<br>• Cerebral infarction: I63<br>• Hypoxic ischemic encephalopathy: P915, P9181, P916 | BORN, MOMBABY and DAD |

| | Description | Applicable codes | Data Sources |
|---|---|---|---|
| | | • Intraventricular hemorrhage (grades 2,3,4): P521, P522<br>• Necrotising enterocolitis: P77<br>• Other respiratory: primary atelectasis respiratory failure: P280, P285<br>• Periventricular leukomalacia: P912<br>• Pneumonia: P23 (preferred code by the Public Health Agency of Canada used), (codes J12 TO J18 used in LAIN et al, not used here)<br>• Respiratory distress syndrome: P220<br>• Seizure: P90, R56<br>• Sepsis/septicaemia (streptococcus, staphylococcus, E.coli, unspecified Gram-negative): P36 (preferred code by the Public Health Agency of Canada used), (codes A40, A415, A419, B951, B962 used in LAIN et al, not used here<br>**CCI Codes**<br>• Any body cavity surgical procedure (substring): 1AA52, 1AA87, 1AC87, 1AE87, 1AF87, 1AG87, 1AJ87, 1AK87, 1AN52, 1AN59, 1AN87, 1AP59, 1AP72, 1AP87, 1AW59, 1AW72, 1AW87, 1AX87, 1BA72, 1BA80, 1BA87, 1BB72, 1BB80, 1BB87, 1BD72, 1BD80, 1BD87, 1BF80, 1BG72, 1BG80, 1BG87, 1BK59, 1BM72, 1BM80, 1BM87, 1BN72, 1BN80, 1BN87, 1BP72, 1BP80, 1BP87, 1BQ72, 1BQ80, 1BQ87, 1BS72, 1BS80, 1BS87, 1BT72, 1BT80, 1BT87, 1GA87, 1GA89, 1GB87, 1GB89, 1GD89, 1GE80, 1GE87, 1GE89, 1GE91, 1GH84, 1GJ86, 1GJ87, 1GK87, 1GK89, 1GM80, 1GM86, 1GM87, 1GN92, 1GR87, 1GR89, 1GR91, | |

| | Description | Applicable codes | Data Sources |
|---|---|---|---|
| | | 1GT78, 1GT87, 1GT89, 1GT91, 1GV87, 1GV89, 1GW87, 1GX80, 1GX86, 1GX87, 1GY70, 1GY72, 1GY86, 1HJ76, 1HJ82, 1HN87, 1HP76, 1HP78, 1HP80, 1HP82, 1HP83, 1HP87, 1HR80, 1HR84, 1HR87, 1HS80 (excl. 1HS80G), 1HS90, 1HT80 (excl. 1HT80G), 1HT89, 1HT90, 1HU80 (excl. 1HU80G), 1HU90, 1HV80 (excl. 1HV80G), 1HV90, 1HW78, 1HW79, 1HX80, 1HX87, 1HX80, 1HZ87, 1IA76, 1IA80, 1IA87, 1IB76, 1IB79, 1IB80, 1IB82, 1IB87, 1IC76, 1IC80, 1IC82, 1IC87, 1ID76, 1ID80, 1ID82, 1ID86, 1ID87, 1IF83, 1IJ76, 1IJ80, 1IM76, 1IM80, 1IM82, 1IM83, 1IM87, 1IN83, 1IN84, 1IN87, 1JE57 (excl. 1JE57G), 1JE76, 1JE80, 1JE87, 1JJ76, 1JJ80, 1JK76, 1JK80, 1JK87, 1JW51 (excL. 1JW51G), 1JW57, 1JW76, 1LA84, 1LC84, 1LD84, 1NA72, 1NA74, 1NA76, 1NA77, 1NA80, 1NA84, 1NA86, 1NA87, 1NA88, 1NA89, 1NA90, 1NA91, 1NA92, 1NE80, 1NF76, 1NF78, 1NF80, 1NF82, 1NF84, 1NF86, 1NF87 (excl. 1NF87B), 1NF89, 1NF90, 1NF91, 1NF92, 1NK76, 1NK77, 1NK80, 1NK82, 1NK84, 1NK87 (excl. 1NK87B), 1NM74, 1NM76, 1NM77, 1NM80, 1NM82, 1NM87 (excl. 1NM87B), 1NM89, 1NM91, 1NP72, 1NP73, 1NP86, 1NQ74 (excl. 1NQ74B), 1NQ80, 1NQ84, 1NQ86, 1NQ87 (excl. 1NQ87B), 1NQ89, 1NQ90, 1NT80, 1NT84, 1NT86, 1NT87, 1NV89, 1OA87, 1OB87, 1OB89, 1OD76, 1OD89, 1OE76, 1OE80, 1OE89, 1OJ76 (excl. 1OJ76B), 1OJ87, 1OJ89, 1OK87, 1OK89, | |

| | Description | Applicable codes | Data Sources |
|---|---|---|---|
| | | 1OK91, 1OT72, 1OT87, 1OT91, 1PB87, 1PB89, 1PC80, 1PC87 (excl. 1PC87D), 1PC89, 1PC91, 1PE57 (excl. 1PE57BD), 1PE80 (excl. 1PE80D), 1PE82, 1PE87 (excl. 1PE87D), 1PE89 (excl. 1PE89D), 1PG76, 1PG77, 1PG80 (excl. 1PG80D), 1PG86, 1PG89, 1PL74 (excl. 1PL74CD), 1PL80, 1PM79, 1PM86, 1PM87 (excl. 1PM87B), 1PM89, 1PM90, 1PM91, 1PM92, 1QE53, 1QE80, 1QE82, 1QE84, 1QE87, 1QE89, 1QG89, 1QM74, 1QM80, 1QM87, 1QM89, 1QM91, 1QN82, 1QT87, 1QT91, 1RB74, 1RB80, 1RB83, 1RB87, 1RB89,1RD89, 1RF51, 1RF72, 1RF74, 1RF80, 1RF87, 1RF89, 1RM87 (excl. 1RM87B), 1RM89, 1RM91, 1RN87, 1RN89, 1RS74, 1RS80, 1RS86, 1RS87, 1RS89, 1RW87, 1RW88, 1RW91, 1RW92, 1SA74, 1SA75, 1SA80, 1SA89, 1SC74, 1SC75, 1SC80, 1SC87, 1SC89, 1SE53, 1SE89 (excl. 1SE89D), 1SF80, 1SF87, 1SF89, 1SG80, 1SG87, 1SH87, 1SM74, 1SM80, 1SM87, 1SN87, 1SN93, 1SQ53, 1SQ74, 1SQ80, 1SQ87, 1SQ91, 1SQ93, 1SW74, 1SY80, 1SY84, 1SY87, 1SZ87, 1VA53, 1VA74, 1VA75, 1VA80, 1VA87, 1VA93, 1VC74, 1VC80, 1VC87, 1VC91, 1VC93, 1VE80, 1VG53, 1VG55, 1VG72, 1VG73, 1VG74, 1VG75, 1VG80, 1VG87, 1VG93, 1VK80, 1VK87, 1VK89, 1VL80, 1VL87, 1VM80, 1VM87, 1VN80, 1VN87, 1VP74, 1VP80, 1VP87, 1VP89, 1VQ74, 1VQ79, 1VQ80, 1VQ82, 1VQ87, 1VQ91, 1VQ93, 1VS72, 1VS80, 1VX87 | |

| | Description | Applicable codes | Data Sources |
|---|---|---|---|
| | | • Any intravenous fluids: 1LZ35CAE6, 1LZ35HAC1, 1LZ35HAC5, 1LZ35HAC6, 1LZ35HAC7, 1LZ35HAE6, 1LZ35HAT7, 1LZ35HAT9, 1LZ35HAZ9, 1LZ35HHC1, 1LZ35HHC5, 1LZ35HHC6, 1LZ35HHC7, 1LZ35HHE0, 1LZ35HHE6, 1LZ35HHT7, 1LZ35HHT9, 1LZ35HHZ9, 1LZ35HRC5, 1LZ35HRC6, 1LZ35HRC7, 1LZ35HRT9, 1LZ35HRZ9<br>• Central venous or arterial catheter: 1KV53HACH, 1KV53HAFT, 1KV53LAFT, 2IM28GP, 2LZ28GQPL, 2LZ28GRPL, 2LZ28JAPL, 1KX53HACH, 1KX53HAFT, 1KX53 LAFT, 2LZ28GQPL, 2LZ28GRPL<br>• Pneumothorax requiring intercostal catheter: 1GV52DA, 1GV52DATS, 1GV52HA, 1GV52HAHE, 1GV52HATK, 1GV52LA, 1GV52LATS, 1GV52LAXXE, 1GV54JATS, 1GV55JATS<br>• Resuscitation: 1HZ30JN, 1HZ30JY, 1GZ30CJ, 1GZ30CJNB, 1GZ30JH<br>• Transfusion of blood or blood products: 1LZ19HHU1A, 1LZ19HHU1J, 1LZ19HHU2A, 1LZ19HHU2J, 1LZ19HHU3J, 1LZ19HHU4J, 1LZ19HHU5J, 1LZ19HHU6A, 1LZ19HHU6J, 1LZ19HHU9A, 1LZ19HHU9J, 1LZ19HMU1, 1LZ19HMU2, 1LZ19HMU9, 1LZ35HAC5<br>• Ventilatory support (mechanical ventilation and/or CPAP): 1GZ31CAEP, 1GZ31CAND, 1GZ31CAPK, 1GZ31CBND, 1GZ31CRND, 1GZ31GPND, | |

| | Description | Applicable codes | Data Sources |
|---|---|---|---|
| | | 1GZ31JAGX, 1GZ31JAMD, 1GZ31JANC, 1GZ31JAPK | |
| **Covariates** | | | |
| **Demographics** | | | |
| **Maternal Age** | | MATAGEATSTILLORLIVEBIRTHYEARS | BORN |
| **Neighborhood Income Quintile** | | (1) Lowest quintile<br>(2) Second quintile<br>(3) Third quintile<br>(4) Fourth quintile<br>(5) Highest quintile<br>If income quintile is unknown → set to (1) lowest | PCCF and CENSUS |
| **Rurality Index** | | (1) Urban (RIO 0-39)<br>(2) Rural (RIO >=40)<br>If rurality is unknown → set to (2) Urban | PCCF and CENSUS |
| **Pre-Pregnancy Conditions** | | | |
| **Obesity** | Yes/No | Pre-pregnancy body mass index (BMI) recorded as BMI ≥30 kg/m² in BORN<br>OR<br>OHIP billing code ICD-9 278 within 2 years prior to estimated date of conception. | BORN and OHIP |
| **Pre-existing Diabetes** | | Incident or Prevalent case in ODD database | ODD |
| **Pre-existing Hypertension** | | Incident or Prevalent case in HYPER database | HYPER |
| **Parity** | Nulliparous/Parous/Grand Multiparous | PARITY<br>Nulliparous if: parity = 0<br>Parous if: parity = 1-5<br>Grand Multiparous = 6+ | BORN |
| **Mental Health Composites** | | | |
| **History of Mental Illness** | A composite measure that includes a diagnosis of a mood or anxiety disorder, psychotic disorder, substance use disorder, self-harm event or other conditions, such as an eating disorder or an | <u>**Emergency Visits and Hospitalizations**</u><br><br>**1. OMHRS/DAD Inpatient hospitalization discharge diagnoses for MHA**<br><br>**From DAD var DX10code1 with the below listed ICD-10-CA codes:**<br>• Include if DX10CODE = F06-F99, <u>OR</u> | OMHRS, DAD, NACRS and OHIP |

| | Description | Applicable codes | Data Sources |
|---|---|---|---|
| | obsessive–compulsive disorder, based on a single emergency department visit or hospital admission, or 2 or more outpatient visits within 2 years of the estimated date of conception. | • DX10CODE2 to DX10CODE10 = X60-X84, Y10-Y19, Y28 AND DX10code1 ne F06-F99<br>o Include visits with suspect diagnosis (suspect = T)<br>**From OMHRS standalone dataset up to March 31, 2016:**<br>• If var AXIS1_DSM4CODE_DISCH1 complete (i.e., listed diagnosis from below present) use it<br>• If not, use PROVDX_DSM4CODE_DISCH1<br>• Exclude OMHRS admission if:<br>o AXIS1_DSM4CODE_DISCH1 in: (290.x OR 294.x) OR<br>o AXIS1_DSM4CODE_DISCH1 is missing AND PROVDX_DSM4CODE_ADM1=2<br><br>**2. NACRS – Emergency department visit diagnoses for MHA**<br><br>**From NACRS var DX10CODE1 with the below listed ICD-10-CA codes**<br>• Include if DX10CODE1 = F06-F99, <u>OR</u><br>• DX10CODE2 to DX10CODE10 = X60-X84, Y10-Y19, Y28 AND DX10CODE1 ne F06-F99<br>o Include visits with suspect diagnoses (suspect = T)<br><br>Substance-Related and Addictive Disorders:<br>• AXIS1_DSM4CODE_DISCH1 = 291.x (all 291 codes, excluding 291.82), 292.x (all 292 codes, excluding 292.85), 303.x (all 303 codes), 304.x (all 304 codes), 305.x (all 305 codes).<br>• PROVDX_DSM4CODE_ADM1 =4<br>• DX10CODE1 = F55, F10 to F19 | |

| | Description | Applicable codes | Data Sources |
|---|---|---|---|
| | | Schizophrenia Spectrum and Other Psychotic Disorders:<br>• AXIS1_DSM4CODE_DISCH1 = 295.x (all 295 codes), 297.x (all 297 codes), 298.x (all 298 codes), 293.81, 293.82<br>• PROVDX_DSM4CODE_ADM1 =5<br>• DX10CODE1 = F06.0, F06.2, F20 (excluding F20.4), F22, F23, F24, F25, F28, F29, F53.1<br><br>Mood and Anxiety Disorders:<br>• AXIS1_DSM4CODE_DISCH1 = 296.x (all 296 codes), 300.4x, 301.13, 311.x, 293.83<br>• PROVDX_DSM4CODE_ADM1 =6<br>• AXIS1_DSM4CODE_DISCH1 = 300.0x, 300.2x, 300.3x, 308.3x, 309.0x, 309.24, 309.28, 309.3x, 309.4x, 309.8x, 309.9x, 293.84<br>• PROVDX_DSM4CODE_ADM1 =7, 15<br>• DX10CODE1 = F06.3, F30, F31, F32, F33, F34, F38, F39, F53.0<br>• DX10CODE1 = F06.4, F40, F41, F42, F43, F48.8, F48.9; F93.1, F93.2<br><br>Deliberate self harm:<br>• DX10CODE2-10 = X60-X84, Y10-Y19, Y28 when DX10CODE1 ne F06-F99<br>• †Deliberate self-harm is an external injury; please specify DXTYPE = alldx or DXTYPE = 9<br>Other<br>• AXIS1_DSM4CODE_DISCH1 = 293.89, 293.90, 300.6, 300.7, 300.8, 300.9, 301, 301.0, 301.2, 301.4, 301.5, 301.6, 301.7, 301.8, 301.9, 307.1, 307.50, 307.51, 307.52, 307.53<br>• PROVDX_DSM4CODE_ADM1 = 12, 16 | |

| | Description | Applicable codes | Data Sources |
|---|---|---|---|
| | | • DX10CODE1 = F06.1, F21, F45, F50, F53.8, F53.9, F60, F61, F69’<br><br>**<u>Outpatient Related Services</u>**<br><br>1. Psychiatrist [SPEC=19] and outpatient (LOCATION: O, L, H) and non-lab service [substr(FEECODE,1,1) ne 'G'] *OR*<br>2. FP/GP [SPEC=00] and MHA diagnosis code [DXCODE] and outpatient (LOCATION: O, L, H) and non-lab service [substr(FEECODE,1,1) ne 'G']<br><br>**Psychotic Disorders**<br>295 Schizophrenia<br>296 Manic-depressive psychoses, involutional melancholia<br>297 Other paranoid states<br>298 Other psychoses<br>**Non-Psychotic Disorders**<br>300 Anxiety neurosis, hysteria, neurasthenia, obsessive-compulsive neurosis, reactive depression<br>301 Personality disorders<br>302 Sexual deviations<br>306 Psychosomatic illness<br>309 Adjustment reaction<br>311 Depressive disorder<br>**Substance Use Disorders**<br>303 Alcoholism<br>304 Drug dependence | |
| **Substance Use** | | | |
| **Alcohol Use in Pregnancy** | Yes/No | PREG_EXPOS_ALCOHOL_ID<br>Yes if:<br>1020440 = More than one drink per week<br>OR<br>1020442 = Episodic excessive drinking (binging) | BORN |

| | Description | Applicable codes | Data Sources |
|---|---|---|---|
| | | No if: any other code, including unknown | |
| **History of Substance Use Disorder** | Substance use components of the above mental illness composite. | | OMHRS, DAD, NACRS and OHIP |
| **Pregnancy Characteristics and Complications** | | | |
| **Conception Type** | Spontaneous/Assisted Reproductive Technology | CONCEPTION_TYPE_ID<br>ART - 1013130, 1013140, 1013170, 1013110, 1013120, 1013150, 3000006<br>Spontaneous or Unknown – 1013160, 1013180 | BORN |
| **Gestational Diabetes** | Yes/No | DIABETES_AND_PREGNANCY_ID<br>=Yes if:<br>1013430 = Gestational diabetes<br>1013440 = Gestational diabetes \ Insulin<br>1013450 = Gestational diabetes \ Insulin \ ACE inhibitors<br>1013460 = Gestational diabetes \ Insulin \ Statins<br>1013465 = Gestational diabetes \ Insulin \ No Ace Inhibitors or Statins<br>1013470 = Gestational diabetes \ No Insulin<br>1013480 = Gestational diabetes \ No Insulin \ No Oral agents<br>1013490 = Gestational diabetes \ No Insulin \ Oral Antihyperglycemic Agents<br>3000001 = Gestational diabetes \ Insulin Status Unknown<br><br>=No if any other code (including unknown or missing) | BORN |
| **Hypertensive Disorder of Pregnancy** | Yes/No | PREG_HYPERTENSION_DISORDER_ID<br>=Yes if:<br>1020800 = Eclampsia<br>1020810 = Gestational Hypertension<br>1020820 = HELLP<br>1020840 = Pre-existing Hypertension with superimposed preeclampsia<br>1020850 = Preeclampsia<br>1020854 = Preeclampsia requiring magnesium sulfate | BORN |

| | Description | Applicable codes | Data Sources |
|---|---|---|---|
| | | =No if any other code (including unknown or missing) | |
| **Placenta Previa** | Yes/No | COMPLICATION_ID<br>=Yes if: 1020340 = Placental \ Placenta previa<br>=No if: Any other code(including unknown or missing) | BORN |
| **Placenta Abruption** | Yes/No | COMPLICATION_ID<br>=Yes if:<br>1020300 = Placental \ Placental Abruption<br>=No if: Any other code (including unknown or missing) | BORN |
| **Delivery Charactersitics** | | | |
| **Gestational Age at Delivery** | Weeks | Gestational Age at Birth (GA_AT_BIRTH_WEEKS)<br>Continuous and according to categories<br>• Extreme Preterm <28 weeks<br>• Very Preterm 28- to <32 weeks<br>• Preterm 32 - 36 weeks | BORN |
| **Delivery Type** | Vaginal<br>C/S | BIRTH_TYPE_ID<br>Vaginal if:<br>1012880 = Assisted Vaginal<br>1012910 = Spontaneous Vaginal<br>1012920 = Vaginal<br>C/S if:<br>1012890 = Induced or Spontaneous Labour Cesarean Section<br>1012900 = No Labour - Cesarean Section<br>Unknown if:<br>1012925 = Unknown | BORN |
| **Type of Labour** | Induced<br>Spontaneous<br>No Labour | LABOUR_TYPE_ID<br>1014630 = Induced<br>1014640 = Spontaneous<br>1014645 = No Labour<br>Note: if unknown, classify as spontaneous | BORN |
| **Birthweight** | Grams | BIRTH_WEIGHT_GRAMS | BORN |
| **Baby Sex** | Female/Male | Sex will come from RPDB sex linked to child's unique identifier (IKN).<br>M = Male<br>F = Female | RPDB and BORN |

| | Description | Applicable codes | Data Sources |
|---|---|---|---|
| | | If missing sex in RPDB, then impute missing value with BORN b_sex variable. | |
| **Jaundice** | Yes/No | BORN (NEONATAL_HEALTH_CONDITIONS_ID)<br>1026460 = Jaundice greater than 475 (Prior to April 2014)<br>1026954 = Hyperbilirubinemia (NICU)<br>1026974 = Hyperbilirubinemia (Prior to April 2018)<br>OR<br>MOMBABY P58 or P59 | BORN and MOMBABY |
| **Maternal readmission** | Readmission to a hospital within 42 days of delivery | | DAD |
| **Infant readmission** | Readmission to a hospital within 42 days of delivery | | DAD |

* For the derived no-substance-exposure indicator, participants classified as unknown across all individual substance fields were included in the no-reported-exposure category, consistent with the prespecified analytic coding approach.

**Supplemental Table 1.** Maternal characteristics by detailed substance exposure groups.

| | Opioid[a] N=4,490 | Opioid & Cannabis[a] N=631 | OAT[a] N=2,339 | OAT & Cannabis[a] N=404 | Opioid & OAT[a] N=771 | Opioid, OAT & Cannabis[a] N=219 | Nicotine Only N=55,829 | Cannabis[a] N=19,091 | Other Substance(s)[b] N=11,449 | No Substance N=864,508 |
|---|---|---|---|---|---|---|---|---|---|---|
| **Maternal Age, yrs** | | | | | | | | | | |
| Mean (SD) | 29.7 (5.5) | 27.3 (5.9) | 28.6 (4.8) | 28.6 (4.8) | 27.9 (5.0) | 28.1 (4.9) | 28.0 (5.7) | 26.2 (5.7) | 29.8 (5.8) | 31.3 (5.0) |
| **Neighborhood Income Quintile, n (%)** | | | | | | | | | | |
| 1 | 1,413 (31.5) | 293 (46.4) | 1,179 (50.4) | 216 (53.5) | 433 (56.2) | 109 (49.8) | 20,383 (36.5) | 7,412 (38.8) | 3,855 (33.7) | 164,127 (19.0) |
| 2 | 906 (20.2) | 135 (21.4) | 443 (18.9) | 96 (23.8) | 125 (16.2) | 48 (21.9) | 13,301 (23.8) | 4,508 (23.6) | 2,312 (20.2) | 168,152 (19.5) |
| 3 | 835 (18.6) | 83 (13.2) | 319 (13.6) | 40 (9.9) | 88 (11.4) | 24 (11.0) | 9,872 (17.7) | 3,148 (16.5) | 2,185 (19.1) | 185,367 (21.4) |
| 4 | 777 (17.3) | 78 (12.4) | 242 (10.3) | 25 (6.2) | 68 (8.8) | 27 (12.3) | 7,419 (13.3) | 2,343 (12.3) | 1,886 (16.5) | 189,953 (22.0) |
| 5 | 559 (12.4) | 42 (6.7) | 156 (6.7) | 27 (6.7) | 57 (7.4) | 11 (5.0) | 4,854 (8.7) | 1,680 (8.8) | 1,211 (10.6) | 156,909 (18.2) |
| **Rurality, n (%)** | | | | | | | | | | |
| Rural | 562 (12.5) | 87 (13.8) | 229 (9.8) | 44 (10.9) | 81 (10.5) | 23 (10.5) | 7,320 (13.1) | 2,247 (11.8) | 821 (7.2) | 56,666 (6.6) |
| Urban | 3,928 (87.5) | 544 (86.2) | 2,110 (90.2) | 360 (89.1) | 690 (89.5) | 196 (89.5) | 48,509 (86.9) | 16,844 (88.2) | 10,628 (92.8) | 807,842 (93.4) |
| **Pre-pregnancy BMI, kg/m$^2$** | | | | | | | | | | |
| Mean (SD) | 26.9 (6.8) | 25.5 (7.0) | 25.4 (6.3) | 24.2 (5.7) | 24.9 (5.7) | 23.8 (6.3) | 26.4 (7.0) | 25.1 (6.8) | 25.5 (6.2) | 25.7 (6.1) |
| Missing Data (%) | 14.0 | 17.0 | 14.7 | 16.6 | 20.5 | 20.1 | 12.0 | 11.9 | 19.4 | 14.4 |
| **Obesity[c], n (%)** | | | | | | | | | | |
| No | 2,796 (62.3) | 420 (66.6) | 1,600 (68.4) | 289 (71.5) | 506 (65.6) | 157 (71.7) | 36,351 (65.1) | 13,416 (70.3) | 7,344 (64.1) | 590,998 (68.4) |
| Yes | 1,083 (24.1) | 106 (16.8) | 400 (17.1) | 49 (12.1) | 107 (13.9) | 19 (8.7) | 12,907 (23.1) | 3,455 (18.1) | 1,943 (17.0) | 153,083 (17.7) |
| Missing | 611 (13.6) | 105 (16.6) | 339 (14.5) | 66 (16.3) | 158 (20.5) | 43 (19.6) | 6,571 (11.8) | 2,220 (11.6) | 2,162 (18.9) | 120,427 (13.9) |
| **Parity, n (%)** | | | | | | | | | | |
| 0 | 1,534 (34.2) | 251 (39.8) | 473 (20.2) | 91 (22.5) | 207 (26.8) | *62-66 | 18,475 (33.1) | 10,331 (54.1) | 4,162 (36.4) | 365,157 (42.2) |
| 1-5 | 2,894 (64.5) | 364 (57.7) | 1,795 (76.7) | 303 (75.0) | 535 (69.4) | 152 (69.4) | 36,589 (65.5) | 8,646 (45.3) | 7,094 (62.0) | 494,232 (57.2) |
| 6+ | 62 (1.4) | 16 (2.5) | 71 (3.0) | 10 (2.5) | 29 (3.8) | *1-5 | 765 (1.4) | 114 (0.6) | 193 (1.7) | 5,119 (0.6) |
| **Comorbidities, n (%)** | | | | | | | | | | |
| Pre-existing Diabetes | 117 (2.6) | 11 (1.7) | 43 (1.8) | 7 (1.7) | 11 (1.4) | *1-5 | 940 (1.7) | 277 (1.5) | 476 (4.2) | 14,195 (1.6) |
| Pre-existing Hypertension | 127 (2.8) | 20 (3.2) | 47 (2.0) | *1-5 | 14 (1.8) | *1-5 | 901 (1.6) | 193 (1.0) | 320 (2.8) | 19,930 (2.3) |
| Hx of Mental Illness[d] | 1,560 (34.7) | 302 (47.9) | 1,856 (79.4) | 343 (84.9) | 572 (74.2) | 169 (77.2) | 14,967 (26.8) | 6,724 (35.2) | 4,649 (40.6) | 109,553 (12.7) |
| Alcohol use in Pregnancy | 30 (0.7) | 18 (2.9) | 10 (0.4) | *1-5 | 14 (1.8) | *1-5 | 312 (0.6) | 225 (1.2) | 401 (3.5) | 448 (0.1) |
| Hx of Substance Use Disorder[d] | 474 (10.6) | 137 (21.7) | 1,737 (74.3) | 320 (79.2) | 512 (66.4) | 151 (68.9) | 2,375 (4.3) | 1,076 (5.6) | 2,678 (23.4) | 4,313 (0.5) |
| Nicotine Exposure | 1,418 (31.6) | 429 (68.0) | 1,703 (72.8) | 352 (87.1) | 595 (77.2) | 191 (87.2) | 55,829 (100.0) | 8,719 (45.7) | 4,579 (40.0) | 0 (0.0) |
| **Pregnancy Complications, n (%)** | | | | | | | | | | |
| Placenta Abruption | 27 (0.6) | *1-5 | 17 (0.7) | 6 (1.5) | 11 (1.4) | *1-5 | 214 (0.4) | 104 (0.5) | 78 (0.7) | 1,919 (0.2) |
| **Maternal Readmission ≥ 42 Days of Delivery, n (%)** | 91 (2.0) | 12 (1.9) | 29 (1.2) | *1-5 | 7 (0.9) | *1-5 | 809 (1.4) | 251 (1.3) | 201 (1.8) | 14,518 (1.7) |

BMI = body mass index; OAT = opioid agonist therapy

[a] Primary exposure categories are mutually exclusive with respect to opioid/OAT/cannabis but may include nicotine.

[b] Patients may use opioid, OAT, cannabis and/or nicotine in addition to the other substance

[c] Obesity is defined as a BMI ≥ 30 kg/m$^2$

[d] History of mental illness and substance use disorder definitions are provided in Appendix 2

* Indicates that the exact values are suppressed due to small numbers. The true number falls within the indicated range of values.

**Supplemental Table 2.** Fetal birth characteristics and neonatal outcomes by detailed substance exposure groups.

| | Opioid[a] | Opioid & Cannabis[a] | OAT[a] | OAT & Cannabis[a] | Opioid & OAT[a] | Opioid, OAT & Cannabis[a] | Nicotine Only | Cannabis[a] | Other Substance(s)[b] | Non-Substance Users |
|---|---|---|---|---|---|---|---|---|---|---|
| | N=4,585 | N=643 | N=2,367 | N=409 | N=778 | N=220 | N=56,628 | N=19,341 | N=11,644 | N=879,090 |
| **Baby Sex, n (%)** | | | | | | | | | | |
| Female | 2,287 (49.9) | 305 (47.4) | 1,169 (49.4) | 204 (49.9) | 346 (44.5) | 115 (52.3) | 28,651 (48.8) | 9,324 (48.2) | 5,711 (49.0) | 428,242 (48.7) |
| Male | 2,298 (50.1) | 338 (52.6) | 1,199 (50.6) | 205 (50.1) | 432 (55.5) | 105 (47.7) | 28,977 (51.2) | 10,023 (51.8) | 5,933 (51.0) | 451,008 (51.3) |
| **Birthweight, g** Mean (SD) | 3210.1 (661.8) | 2959.7 (685.0) | 3141.1 (651.2) | 2898.5 (671.3) | 3126.6 (689.4) | 2947.8 (636.0) | 3197.8 (598.5) | 3110.3 (647.2) | 3137.9 (646.6) | 3348.8 (591.6) |
| Female | 3157.2 (650.8) | 2983.7 (630.6) | 3070.7 (637.9) | 2829.7 (659.0) | 3020.7 (647.5) | 2880.9 (584.6) | 3134.2 (580.5) | 3057.6 (627.8) | 3083.5 (630.3) | 3289.5 (574.5) |
| Male | 3262.8 (668.5) | 2938.0 (730.9) | 3209.6 (656.9) | 2966.9 (677.9) | 3211.4 (710.6) | 3020.9 (683.3) | 3258.5 (608.9) | 3159.3 (661.0) | 3190.3 (657.7) | 3405.0 (601.9) |
| **Gestational Age, weeks** Mean (SD) | 38.0 (2.4) | 37.6 (2.8) | 38.1 (2.4) | 37.6 (2.9) | 38.1 (2.6) | 37.8 (2.6) | 38.5 (2.1) | 38.3 (2.5) | 38.1 (2.4) | 38.7 (2.0) |
| **Stillbirth, n (%)** | 17 (0.4) | *1-5 | 12 (0.5) | 0 (0.0) | 7 (0.9) | *1-5 | 160 (0.3) | 70 (0.4) | 32 (0.3) | 1,502 (0.2) |
| **Small for Gestational Age, n (%)** | | | | | | | | | | |
| <3rd %ile | 129 (2.8) | 48 (7.5) | 113 (4.8) | 35 (8.6) | 50 (6.4) | 16 (7.3) | 2,656 (4.7) | 1,131 (5.8) | 569 (4.9) | 19,729 (2.2) |
| <10th %ile | 491 (10.7) | 128 (19.9) | 362 (15.3) | 98 (24.0) | 145 (18.6) | 51 (23.2) | 8,510 (15.0) | 3,543 (18.3) | 1,779 (15.3) | 79,082 (9.0) |
| **IUGR[c], n (%)** | 120 (2.6) | 35 (5.4) | 103 (4.4) | 21 (5.1) | 35 (4.5) | 13 (5.9) | 2,022 (3.6) | 1,014 (5.2) | 443 (3.8) | 17,052 (1.9) |
| **Severe Neonatal Morbidity, n (%)** | 558 (12.2) | 109 (17.0) | 298 (12.6) | 64 (15.6) | 105 (13.5) | 34 (15.5) | 4,567 (8.1) | 2,055 (10.6) | 1,257 (10.8) | 63,039 (7.2) |
| **Neonatal Abstinence Syndrome, n (%)** | 820 (17.9) | 206 (32.0) | 1,596 (67.4) | 303 (74.1) | 525 (67.5) | 165 (75.0) | 720 (1.3) | 546 (2.8) | 2,493 (21.4) | 1,012 (0.1) |
| **Jaundice, n (%)** | 772 (16.8) | 129 (20.1) | 589 (24.9) | 103 (25.2) | 204 (26.2) | 66 (30.0) | 5,241 (9.3) | 2,395 (12.4) | 1,530 (13.1) | 80,777 (9.2) |
| **Infant Readmission ≤ 42 Days of Delivery, n (%)** | 423 (9.2) | 69 (10.7) | 234 (9.9) | 50 (12.2) | 101 (13.0) | 20 (9.1) | 3,611 (6.4) | 1,483 (7.7) | 1,026 (8.8) | 59,388 (6.8) |
| **Neonatal Mortality, n (%)** | 26 (0.6) | *1-5 | 19 (0.8) | *1-5 | 13 (1.7) | *1-5 | 313 (0.6) | 160 (0.8) | 70 (0.6) | 3,633 (0.4) |

Primary exposure categories are mutually exclusive with respect to opioid/OAT/cannabis but may include nicotine. BMI = body mass index; OAT = opioid agonist therapy

OAT = opioid agonist therapy; IUGR = intrauterine growth restriction

[a] Patients may use nicotine in addition to the substances listed

[b] Patients may use opioid, OAT, cannabis and/or nicotine in addition to the other substance

[c] IUGR was based on clinician diagnosis in BORN Ontario.

* Indicates that the exact values are suppressed due to small numbers. The true number falls within the indicated range of values.

**Supplemental Table 3.** Incidence and relative risk (95% confidence intervals) for outcomes when opioid, OAT, and cannabis exposures are modelled independently.

| | | Incidence per 1000 Pregnancies (95%CI) | Unadjusted RR (95%CI) | Adjusted Model 1[a] RR (95%CI) | Adjusted Model 2[b] RR (95%CI) | Interaction with Cannabis, p-value |
|---|---|---|---|---|---|---|
| **Outcome: IUGR[c]** | | | | | | |
| Opioid | Yes | 38.3 (34.1-42.9) | 1.69 (1.50 - 1.91) | 1.07 (0.94 - 1.21) | 1.15 (1.00 - 1.33) | 0.12 |
| | No | 21.2 (21.0-21.5) | 1.00 (Ref) | 1.00 (Ref) | 1.00 (Ref) | |
| OAT | Yes | 50.3 (44.5-56.6) | 2.33 (2.06 - 2.64) | 1.35 (1.18 - 1.55) | 1.47 (1.26 - 1.71) | 0.049 |
| | No | 21.2 (20.9-21.5) | 1.00 (Ref) | 1.00 (Ref) | 1.00 (Ref) | |
| Cannabis | Yes | 53.8 (50.9-56.9) | 2.45 (2.31 - 2.61) | 1.72 (1.61 - 1.83) | 1.77 (1.66 - 1.89) | - |
| | No | 20.6 (20.3-20.9) | 1.00 (Ref) | 1.00 (Ref) | 1.00 (Ref) | |
| **Outcome: SGA <3%ile** | | | | | | |
| Opioid | Yes | 46.5 (41.8-51.5) | 1.83 (1.65 - 2.03) | 1.06 (0.95 - 1.18) | 1.11 (0.98 - 1.26) | 0.19 |
| | No | 24.9 (24.6-25.2) | 1.00 (Ref) | 1.00 (Ref) | 1.00 (Ref) | |
| OAT | Yes | 61.4 (54.9-68.3) | 2.43 (2.18 - 2.71) | 1.28 (1.14 - 1.44) | 1.31 (1.14 - 1.50) | 0.59 |
| | No | 24.9 (24.6-25.2) | 1.00 (Ref) | 1.00 (Ref) | 1.00 (Ref) | |
| Cannabis | Yes | 61.5 (58.3-64.7) | 2.45 (2.32 - 2.59) | 1.52 (1.43 - 1.61) | 1.54 (1.45 - 1.64) | - |
| | No | 24.2 (23.9-24.5) | 1.00 (Ref) | 1.00 (Ref) | 1.00 (Ref) | |
| **Outcome: SGA <10%ile** | | | | | | |
| Opioid | Yes | 143.2 (135.0-151.8) | 1.45 (1.37 - 1.53) | 1.02 (0.96 - 1.08) | 1.06 (0.99 - 1.13) | 0.041 |
| | No | 96.2 (95.5-96.8) | 1.00 (Ref) | 1.00 (Ref) | 1.00 (Ref) | |
| OAT | Yes | 181.6 (170.5-193.4) | 1.84 (1.74 - 1.96) | 1.22 (1.14 - 1.30) | 1.23 (1.14 - 1.32) | 0.64 |
| | No | 96.1 (95.4-96.7) | 1.00 (Ref) | 1.00 (Ref) | 1.00 (Ref) | |
| Cannabis | Yes | 187.7 (182.2-193.4) | 1.89 (1.84 - 1.95) | 1.38 (1.34 - 1.42) | 1.39 (1.35 - 1.44) | - |
| | No | 94.3 (93.7-94.9) | 1.00 (Ref) | 1.00 (Ref) | 1.00 (Ref) | |
| **Outcome: SNM** | | | | | | |
| Opioid | Yes | 138.7 (130.6-147.2) | 1.82 (1.71 - 1.93) | 1.46 (1.37 - 1.55) | 1.51 (1.41 - 1.62) | 0.034 |
| | No | 73.4 (72.8-73.9) | 1.00 (Ref) | 1.00 (Ref) | 1.00 (Ref) | |
| OAT | Yes | 141.9 (132.1-152.3) | 1.89 (1.76 - 2.03) | 1.54 (1.42 - 1.66) | 1.58 (1.45 - 1.73) | 0.21 |
| | No | 73.5 (73.0-74.0) | 1.00 (Ref) | 1.00 (Ref) | 1.00 (Ref) | |
| Cannabis | Yes | 113.9 (109.6-118.4) | 1.54 (1.48 - 1.60) | 1.33 (1.27 - 1.38) | 1.35 (1.30 - 1.41) | - |
| | No | 72.9 (72.4-73.4) | 1.00 (Ref) | 1.00 (Ref) | 1.00 (Ref) | |

OAT = opioid agonist therapy; IUGR = intrauterine growth restriction; SGA = small for gestational age; SNM = severe neonatal morbidity

[a]Model 1 - adjusted for maternal age, income, rurality, obesity, pre-existing diabetes and hypertension, parity, nicotine exposure, other substance use, singleton or multiple, and fetal sex. [b]Model 2 – adjusted for all Model 1 covariates and additionally included opioid × cannabis and OAT × cannabis interaction terms.

**Supplemental Table 4.** Incidence and relative risk (95% confidence intervals) for outcomes models stratified by cannabis exposure.

| | | Cannabis Exposed | | | Cannabis Unexposed | | |
|---|---|---|---|---|---|---|---|
| | | Incidence per 1000 Pregnancies (95%CI) | Unadjusted RR (95%CI) | Adjusted RR[a] (95%CI) | Incidence per 1000 Pregnancies (95%CI) | Unadjusted RR (95%CI) | Adjusted RR[a] (95%CI) |
| **Outcome: IUGR[b]** | | | | | | | |
| Opioid | Yes | 60.0 (48.2-73.7) | 1.08 (0.86 - 1.35) | 0.96 (0.76 - 1.22) | 33.2 (28.8-38.0) | 1.54 (1.34 - 1.78) | 1.14 (0.99 - 1.32) |
| | No | 53.4 (50.4-56.6) | 1.00 (Ref) | 1.00 (Ref) | 20.5 (20.2-20.8) | 1.00 (Ref) | 1.00 (Ref) |
| OAT | Yes | 64.8 (50.5-81.9) | 1.20 (0.95 - 1.53) | 1.10 (0.85 - 1.43) | 46.6 (40.4-53.5) | 2.26 (1.96 - 2.61) | 1.45 (1.24 - 1.69) |
| | No | 53.3 (50.3-56.4) | 1.00 (Ref) | 1.00 (Ref) | 20.5 (20.2-20.8) | 1.00 (Ref) | 1.00 (Ref) |
| **Outcome: SGA <3%ile** | | | | | | | |
| Opioid | Yes | 74.6 (61.4 -89.8) | 1.23 (1.01 - 1.49) | 0.99 (0.79 - 1.22) | 39.8 (35.0-45.0) | 1.64 (1.45 - 1.85) | 1.09 (0.97 - 1.24) |
| | No | 60.6 (57.3-63.9) | 1.00 (Ref) | 1.00 (Ref) | 24.1 (23.8-24.4) | 1.00 (Ref) | 1.00 (Ref) |
| OAT | Yes | 87.0 (70.3-106.5) | 1.46 (1.20 - 1.79) | 1.27 (1.01 - 1.59) | 54.9 (48.2-62.4) | 2.26 (1.98 - 2.57) | 1.26 (1.10 - 1.45) |
| | No | 60.2(57.1-63.5) | 1.00 (Ref) | 1.00 (Ref) | 24.1 (23.7-24.4) | 1.00 (Ref) | 1.00 (Ref) |
| **Outcome: SGA <10%ile** | | | | | | | |
| Opioid | Yes | 208.5 (186.1-233.0) | 1.12 (1.01 - 1.24) | 0.96 (0.86 - 1.07) | 127.7 (119.1-136.8) | 1.34 (1.25 - 1.43) | 1.05 (0.98 - 1.12) |
| | No | 186.3 (180.6-192.1) | 1.00 (Ref) | 1.00 (Ref) | 94.1 (93.5-94.7) | 1.00 (Ref) | 1.00 (Ref) |
| OAT | Yes | 243.5 (215.0-274.8) | 1.33 (1.19 - 1.49) | 1.20 (1.06 - 1.36) | 166.2 (154.3-178.8) | 1.74 (1.62 - 1.87) | 1.20 (1.11 - 1.29) |
| | No | 185.0 (179.4-190.8) | 1.00 (Ref) | 1.00 (Ref) | 94.0 (93.4-94.6) | 1.00 (Ref) | 1.00 (Ref) |
| **Outcome: SNM** | | | | | | | |
| Opioid | Yes | 165.9 (145.9-187.8) | 1.50 (1.33 - 1.70) | 1.25 (1.09 - 1.43) | 132.3 (123.5-141.6) | 1.75 (1.64 - 1.88) | 1.52 (1.41 - 1.63) |
| | No | 110.4 (106.0-114.9) | 1.00 (Ref) | 1.00 (Ref) | 72.5 (72.0-73.1) | 1.00 (Ref) | 1.00 (Ref) |
| OAT | Yes | 166.7 (143.2-192.9) | 1.47 (1.27 - 1.69) | 1.31 (1.12 - 1.53) | 135.7 (125.0-147.2) | 1.84 (1.70 - 1.99) | 1.59 (1.46 - 1.73) |
| | No | 111.4 (107.0-115.9) | 1.00 (Ref) | 1.00 (Ref) | 72.6 (72.1-73.2) | 1.00 (Ref) | 1.00 (Ref) |

OAT = opioid agonist therapy; IUGR = intrauterine growth restriction; SGA = small for gestational age; SNM = severe neonatal morbidity

[a]Model adjusted for maternal age, income, rurality, obesity, pre-existing diabetes and hypertension, parity, nicotine exposure, other substance use, singleton or multiple, and fetal sex.

[b] IUGR was based on clinician diagnosis in BORN Ontario.
